# Personal night light exposure predicts incidence of cardiovascular diseases in >88,000 individuals

**DOI:** 10.1101/2025.06.20.25329961

**Authors:** Daniel P. Windred, Angus C. Burns, Martin K. Rutter, Jacqueline M. Lane, Richa Saxena, Frank A. J. L. Scheer, Sean W. Cain, Andrew J. K. Phillips

**Affiliations:** Flinders Health and Medical Research Institute (Sleep Health), Flinders University, Bedford Park, SA, Australia; Division of Sleep and Circadian Disorders, Brigham and Women’s Hospital, Boston, MA, USA; Division of Sleep Medicine, Harvard Medical School, Boston, MA, USA; Program in Medical and Population Genetics, Broad Institute, Cambridge, MA, USA; Center for Genomic Medicine, Massachusetts General Hospital, Boston, MA, USA; Division of Diabetes, Endocrinology and Gastroenterology, School of Medical Sciences, University of Manchester, Manchester, UK; Diabetes, Endocrinology and Metabolism Centre, Manchester University NHS Foundation Trust, Manchester Academic Health Science Centre, NIHR Manchester Biomedical Research Centre, Manchester, UK; Department of Anesthesia, Critical Care and Pain Medicine, Massachusetts General Hospital and Harvard Medical School, Boston, MA, USA

## Abstract

**Importance:** Light at night causes circadian disruption, which is a known risk factor for adverse cardiovascular outcomes. However, it is not well understood whether personal light exposure patterns predicts and individual’s risk of cardiovascular diseases.

**Objective:** To assess whether day and night light exposure predicts incidence of cardiovascular diseases, and whether relationships of light with cardiovascular diseases differ according to genetic susceptibility, sex, and age.

**Design:** Prospective cohort study.

**Setting:** United Kingdom.

**Participants:** N=88,905, age (M±SD) = 62.4±7.8 years, 56.9% female.

**Exposure:** Approximately 13 million hours of personal light exposure data, tracked by wrist-worn light sensors (one week each).

**Main Outcomes and Measures:** Incidence of coronary artery disease, myocardial infarction, heart failure, atrial fibrillation, and stroke, derived from UK National Health Service records, across a 9.5-year follow-up after light tracking.

**Results:** People with the brightest nights (90-100^th^ percentiles) had significantly higher risks of developing coronary artery disease (adjusted-HR range: 1.23-1.32), myocardial infarction (aHRs: 1.42-1.47), heart failure (aHRs: 1.45-1.56), atrial fibrillation (aHRs: 1.28-1.32), and stroke (aHRs: 1.28-1.30), compared to people with dark nights (0-50^th^ percentiles). These relationships were robust after adjusting for established risk factors for cardiovascular health, including physical activity, smoking, alcohol, diet, sleep duration, socioeconomic status, and polygenic risk. Relationships of night light with risk of heart failure and coronary artery disease were stronger for women, and relationships of night light with risk of heart failure and atrial fibrillation were stronger for younger individuals in this cohort.

**Conclusions and Relevance:** Night light exposure was a significant risk factor for developing cardiovascular diseases in this cohort. In addition to current preventative measures, avoiding light at night may be a useful strategy for reducing risks of cardiovascular diseases.

**Key Points:** 

**Question:** Does personal light exposure predict incidence of cardiovascular diseases?

**Findings:** In this study of ∼89,000 adults aged >40 years, exposure to brighter light at night predicted higher incidence of coronary artery disease, myocardial infarction, heart failure, atrial fibrillation, and stroke, independent of established cardiovascular risk factors.

**Meaning:** Avoiding exposure to night light may lower risk for cardiovascular diseases.

## Introduction

Robust circadian rhythms are vital for healthy cardiovascular function. Circadian rhythms have been observed in systolic and diastolic blood pressure,^1,2^ platelet activation,^3^ fibrinolysis;^4^ vascular endothelial function;^5^ circulating cortisol, epinephrine, and norepinephrine;^2^ glucose tolerance;^6^ and heart rate average, heart rate variability, QT interval, and PR segment.^1,7,8^ Short-term circadian disruption in humans causes hypercoagulability,^9^ elevated heart rate,^10^ elevated blood pressure, inflammation, and reduced cardiac vagal modulation.^11,12^ Long-term circadian disruption in animal models causes myocardial fibrosis, hypertrophy, impaired contractility, adverse cardiac remodeling, and accelerated progression to heart failure.^13–15^ Epidemiological evidence demonstrates higher risks of adverse cardiovascular events, coronary heart disease, heart failure, atrial fibrillation, and mortality due to cardiovascular disease in rotating shift workers^16–20^ who have long-term exposure to circadian disruption.

Light at night causes circadian disruption,^21–23^ and is therefore a potential determinant of cardiovascular disease risk. Higher risks for coronary artery disease^24^ and stroke^25^ have been observed in people living in urban environments with brighter outdoor night light, as measured by satellite. Brighter night light has been cross-sectionally related to atherosclerosis,^26,27^ obesity, hypertension, and diabetes^28^ in small but well-characterized cohorts, using bedroom^26,27^ and wrist-worn^28^ light sensors. Moreover, experimental exposure to night light elevates heart rate and alters sympathovagal balance.^29^ However, current evidence linking night light with cardiovascular risk is mostly within small cohorts, or relies on geospatial-level measurements of outdoor lighting, rather than measures of personal light exposure.^30,31^

Using data captured from wrist-worn light sensors, in a large cohort of ∼89,000 individuals, we recently observed higher risk of mortality by cardiometabolic causes in those exposed to brighter nights and darker days.^32^ In the same cohort, brighter nights also predicted higher incidence of type 2 diabetes,^33^ an established risk factor for cardiovascular diseases.^34^

We therefore assessed whether personal day and night light exposure predicted incident coronary artery disease, myocardial infarction, heart failure, atrial fibrillation, and stroke, over 9.5-years of follow-up in UK Biobank participants.

## Methods

### Overview

Approximately 502,000 UK Biobank participants were recruited between 2006-2010, and 103,669 participants wore light-tracking devices (Axivity AX3, Newcastle upon Tyne, United Kingdom; peak spectral sensitivity 560 nm) on their dominant wrist for one week between 2013 and 2016. Incident cardiovascular diseases were captured up to November 2022. Detailed information on the data collection protocol is available on the UK Biobank website (see S1). Ethical approval was granted by the North West Multi-centre Research Ethics Committee.

### Exposure: personal light tracking

Extraction of personal day and night light exposures from wrist-worn light sensor data in this cohort has been previously reported.^32,33,35^ In short, sensor data (100Hz) were down-sampled, cleaned for periods of non-wear and data corruption, and transformed according to the device manual and subsequent testing under reference lighting conditions. Data were then grouped into 24-hour light exposure profiles for each participant, represented by average light exposure within each of 48 half-hour clock time intervals (e.g., all light between 00:00-00:30). Factor analysis was applied to the 24-hour light exposure profiles, extracting day (07:30–20:30) and night (00:30–06:00) light exposure times (see S2).

### Outcome: incident cardiovascular diseases

Diagnoses of cardiovascular diseases were derived from hospital admissions, primary care, self-report, and death register records, according to ICD-9 and ICD-10 diagnostic criteria. Myocardial infarction and stroke were defined according to the UK Biobank’s algorithmically-defined outcomes. Myocardial infarction captured both ST-segment-elevated and non-ST-segment-elevated events. Stroke captured ischemic stroke, intracerebral hemorrhage, and subarachnoid hemorrhage. ICD codes included in myocardial infarction and stroke definitions are documented on the UK Biobank website (see S1). Coronary artery disease captured acute and chronic ischemic heart disease, myocardial infarction, and coronary artery operations (see S2). Atrial fibrillation was defined according to the first occurrence of ICD-10 code I48, or operation for atrial fibrillation (see S2). Heart failure was defined according to the first occurrence of ICD-10 code I50. Participants with each cardiovascular outcome prior to light tracking were excluded from analyses (Figure 1).

**Figure 1.**
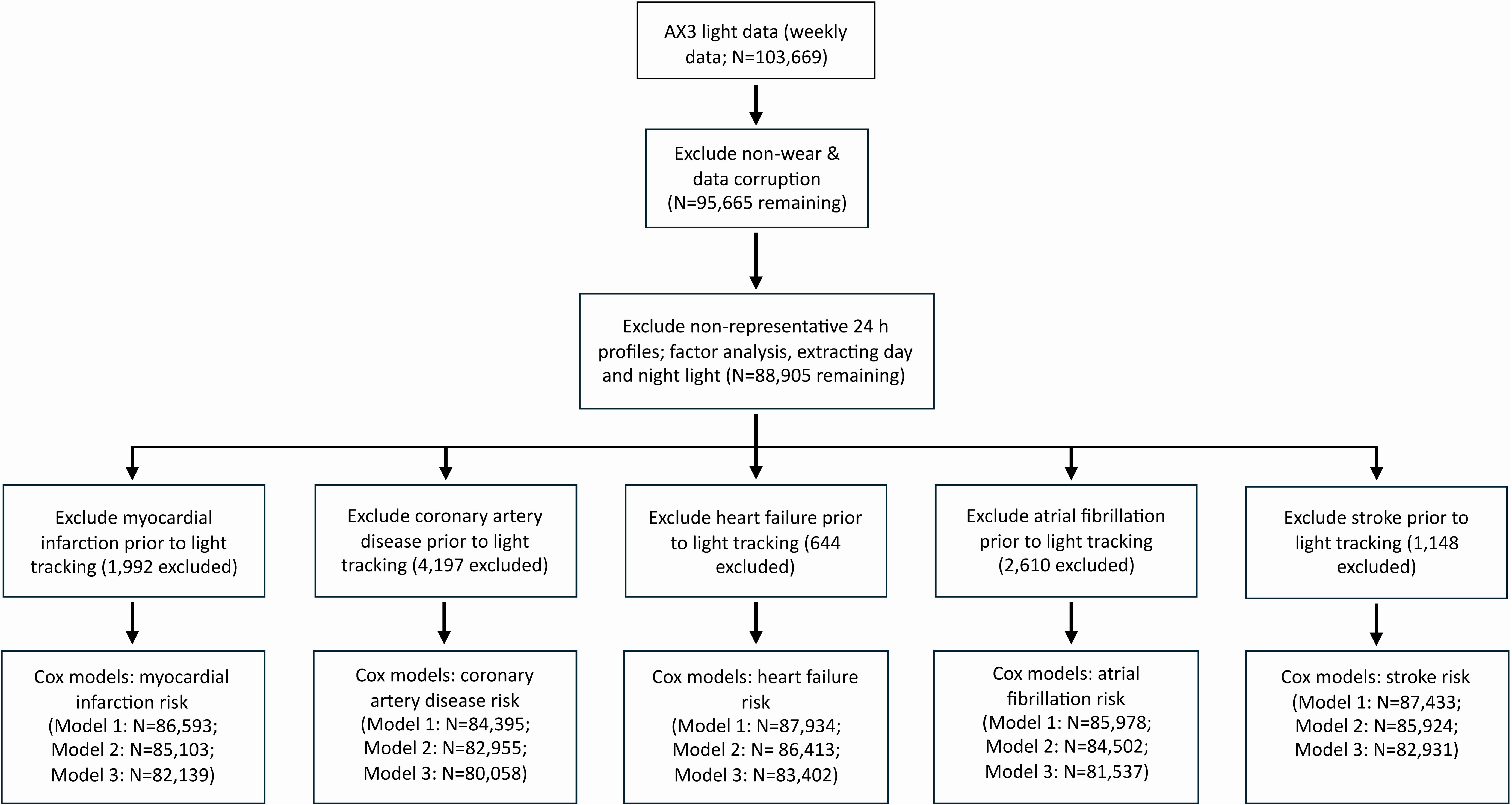
Consort diagram detailing participant-level exclusions between data collection and final analyses.

### Covariates

Participant age, sex, ethnicity, yearly household income, education level, employment status, material deprivation (Townsend Deprivation Index),^36^ urbanicity of residential location, alcohol consumption (days per week), smoking status (never/previous/current), healthy diet score,^37^ and shift work status were derived from questionnaires administered at a baseline assessment centre visit, between 2006 and 2010. Photoperiod was defined as the time between sunrise and sunset during light tracking. Average physical activity was estimated from weekly accelerometer recordings.^38^ Pre-existing diabetes was defined as occurrence of ICD-10 codes E10-11, or self-reported diagnosis, prior to light tracking. Pre-existing hypertension was defined as the occurrence of ICD-10 codes I10-13, or I15, or measured hypertension at baseline physical assessment, prior to light tracking. Body mass index (BMI) and cholesterol ratio were derived from physical measurements collected during baseline assessment. Sleep duration and sleep efficiency were estimated from weekly accelerometer recordings using a validated sleep-wake estimation method (GGIR), as reported previously.^39–41^ Polygenic risk scores for coronary artery disease,^42^ myocardial infarction,^42^ heart failure,^43^ atrial fibrillation,^42^ and stroke^44^ were generated using PRS-continuous shrinkage,^45^ and scored in the UK Biobank actigraphy cohort using PLINK 2^46^ (see S2). A detailed description of each covariate is provided in Table S3.

### Statistical analysis

Risks of incident cardiovascular diseases were assessed using Cox proportional hazards models, including day and night light as model predictors (see S2). Day and night light were split into 0-50^th^, 50-70^th^, 70-90^th^, and 90-100^th^ light exposure percentiles for inclusion in Cox models (see S2). The 0-50^th^ percentile groups were set as referent groups, representing participants in the darkest environments. Time since light tracking was used as the timescale in all models. Data were right-censored at the end of the observation period (29^th^ November 2022), or at participant mortality if this occurred earlier.

Primary models were adjusted at three levels: Model 1 adjusted for age, sex, ethnicity, and photoperiod; Model 2 additionally adjusted for education, employment, income, and deprivation; and Model 3 further adjusted for physical activity, smoking status, alcohol consumption, diet, and urbanicity. Supplementary models included separate adjustments of Model 3 for pre-existing diabetes, hypertension, high BMI, high cholesterol ratio, short, long, or inefficient sleep, and exclusion of shift workers (S5-10). Selected covariates were potential confounders of relationships between light exposure and cardiovascular risks. Covariates in Model 3 and supplementary models were also potentially indirect or direct mediators of relationships between light exposure and cardiovascular risks.

Interactions of night light exposure with age, sex, and polygenic risk were assessed, in additional Cox models predicting each cardiovascular outcome (see S2). First, dose-response relationships of night light with risk of each cardiovascular outcome were assessed by including night light as a log-linear predictor in Model 3. Subsequently, interactions of age and sex with log-linear night light were added to Model 3. Additionally, an interaction of polygenic risk score with log-linear night light was added to Model 3. Models including polygenic risk scores were restricted to individuals of European ancestry and adjusted for the top 5 principal components of ancestry, to control for potential residual population stratification within the European ancestry sub-population (see S2).

## Results

Participants included in the analyses (N=88,905) had light data across all clock times and were free of each cardiovascular outcome at the time of light tracking (Figure 1). Follow-up time between light tracking and the end of follow-up (November 2022 or participant mortality) averaged 7.9±1.0 years. The interval between the first light recording and the end of follow-up was 9.5 years. Numbers of cases of each cardiovascular outcome, and participant characteristics split by light exposure percentiles, are provided in Table 1. Participant characteristics for the total sample with complete light data, and for sub-groups without each cardiovascular outcome prior to light tracking, are provided in Table S4.

**Table 1.** Participant characteristics and proportions with incident cardiovascular diseases, by light exposure percentiles, by day and night.

|  | Night light exposure percentile |  |  |  | Day light exposure percentile |  |  |  |
| --- | --- | --- | --- | --- | --- | --- | --- | --- |
|  | 0-50% | 50-70% | 70-90% | 90-100% | 0-50% | 50-70% | 70-90% | 90-100% |
| **Age** |  |  |  |  |  |  |  |  |
| Mean±SD | 62.8±7.9 | 61.8±7.9 | 62.0±7.8 | 62.4±7.7 | 62.1±8.0 | 62.4±7.8 | 62.6±7.7 | 63.5±7.4 |
| Range | 43.5 to 78.9 | 43.5 to 79.0 | 43.7 to 78.8 | 43.8 to 78.4 | 43.6 to 79.0 | 43.6 to 78.7 | 43.5 to 78.8 | 43.5 to 78.5 |
| **Sex** (% male, N) | 41.3 (18353) | 45.0 (7998) | 44.7 (7949) | 45.2 (4022) | 42.2 (18774) | 42.1 (7478) | 43.4 (7719) | 48.9 (4351) |
| **Ethnicity** (% white, N) | 97.7 (43296) | 96.7 (17133) | 96.2 (17033) | 95.6 (8462) | 96.3 (42669) | 97.0 (17190) | 97.7 (17315) | 98.7 (8750) |
| **Employment status** (% employed, N) | 59.7 (26360) | 64.5 (11388) | 65.0 (11474) | 63.7 (5622) | 63.9 (28201) | 61.8 (10903) | 60.7 (10720) | 56.9 (5020) |
| **Income** |  |  |  |  |  |  |  |  |
| % <£18k, N | 12.9 (5679) | 13.1 (2307) | 12.9 (2278) | 14.0 (1238) | 13.5 (5959) | 12.9 (2283) | 12.6 (2221) | 11.7 (1039) |
| % £18-29.9k, N | 22.1 (9753) | 21.4 (3779) | 21.2 (3747) | 21.8 (1924) | 21.6 (9537) | 22.0 (3884) | 21.6 (3815) | 22.2 (1967) |
| % £30-51.9k, N | 25.9 (11452) | 26.3 (4637) | 26.0 (4592) | 25.3 (2231) | 25.9 (11431) | 25.4 (4489) | 26.4 (4662) | 26.3 (2330) |
| % £52-100k, N | 22.7 (10016) | 23.1 (4073) | 23.4 (4129) | 22.2 (1957) | 22.8 (10058) | 22.8 (4017) | 23.1 (4079) | 22.8 (2021) |
| % >£100k, N | 6.3 (2767) | 6.9 (1219) | 7.1 (1261) | 7.3 (644) | 6.6 (2917) | 6.5 (1147) | 6.9 (1220) | 6.9 (607) |
| **Education** |  |  |  |  |  |  |  |  |
| % other, N | 48.1 (21144) | 48.4 (8529) | 48.6 (8547) | 48.9 (4300) | 47.8 (21013) | 48.7 (8552) | 48.7 (8577) | 49.6 (4378) |
| % university/college, N | 43.4 (19105) | 43.4 (7639) | 43.5 (7657) | 42.9 (3775) | 44.0 (19367) | 43.2 (7596) | 43.0 (7566) | 41.4 (3647) |
| **Townsend Deprivation Index** |  |  |  |  |  |  |  |  |
| M±SD | -1.92±2.70 | -1.69±2.82 | -1.61±2.88 | -1.39±2.99 | -1.58±2.90 | -1.78±2.76 | -1.92±2.69 | -2.25±2.44 |
| Range | -6.26 to 10.50 | -6.26 to 9.89 | -6.26 to 9.99 | -6.26 to 9.89 | -6.26 to 10.50 | -6.26 to 9.89 | -6.26 to 9.89 | -6.26 to 8.94 |
| **Smoking** |  |  |  |  |  |  |  |  |
| % previous, N | 34.8 (15409) | 36.1 (6391) | 37.9 (6725) | 38.8 (3439) | 35.2 (15579) | 36.1 (6396) | 36.9 (6539) | 38.9 (3450) |
| % current, N | 5.4 (2411) | 7.3 (1288) | 8.2 (1451) | 10.2 (907) | 7.1 (3130) | 7.0 (1247) | 6.6 (1167) | 5.8 (513) |
| **Alcohol** (Mean±SD, days per week) | 2.97±2.48 | 2.98±2.50 | 2.99±2.51 | 2.94±2.55 | 2.87±2.48 | 2.98±2.50 | 3.07±2.51 | 3.29±2.53 |
| **Urbanicity** (% >10,000 population, N) | 83.2 (36620) | 83.7 (14741) | 85.6 (15075) | 86.5 (7598) | 85.5 (37603) | 84.1 (14799) | 83.1 (14628) | 79.4 (7004) |
| **Physical activity** |  |  |  |  |  |  |  |  |
| M±SD | 27.9±7.9 | 28.6±8.3 | 28.5±8.2 | 27.8±8.3 | 27.3±7.9 | 28.2±8.0 | 29±8.1 | 30.6±8.5 |
| Range | 4.8 to 69.2 | 6.5 to 69.3 | 5.9 to 69.2 | 5.1 to 69.4 | 5.1 to 69.3 | 4.9 to 67.9 | 6.5 to 69.4 | 4.8 to 67.4 |
| **Diet score** (% healthy, N) | 26.6 (11476) | 25.2 (4362) | 24.9 (4298) | 24.7 (2123) | 25.3 (10920) | 25.8 (4468) | 26.2 (4530) | 26.9 (2341) |
| **Cardiovascular cases** |  |  |  |  |  |  |  |  |
| Coronary artery disease (% , N) | 6.48 (2817) | 6.92 (1202) | 7.29 (1265) | 7.84 (678) | 6.73 (2921) | 6.81 (1186) | 6.92 (1204) | 7.50 (651) |
| Myocardial infarction (% , N) | 1.78 (773) | 2.06 (358) | 2.18 (379) | 2.53 (219) | 1.93 (838) | 1.92 (334) | 2.00 (348) | 2.41 (209) |
| Heart failure (% , N) | 2.11 (917) | 2.30 (400) | 2.27 (395) | 2.86 (247) | 2.28 (989) | 2.33 (406) | 2.09 (364) | 2.31 (200) |
| Atrial fibrillation (% , N) | 6.45 (2805) | 6.55 (1138) | 6.93 (1204) | 7.90 (683) | 6.54 (2839) | 6.83 (1189) | 6.51 (1132) | 7.72 (670) |
| Stroke (% , N) | 2.43 (1058) | 2.45 (425) | 2.53 (440) | 2.79 (241) | 2.56 (1110) | 2.44 (424) | 2.47 (430) | 2.31 (200) |
| **Light exposure (lux)** |  |  |  |  |  |  |  |  |
| Median (IQR) | 0.62 (0.49-0.80) | 2.48 (1.64-3.93) | 16.4 (10.1-27.2) | 105 (69.4-191) | 426 (221-675) | 1320 (1150-1520) | 2310 (2000-2680) | 3810 (3450-4350) |
| Range | 0 - 1.21 | 1.21 - 6.28 | 6.28 - 48.3 | 48.3 - 6400 | 4.29 - 991 | 991 - 1750 | 1750 - 3140 | 3140 - 7890 |
| **Photoperiod (range:7.5-17.0 h)** |  |  |  |  |  |  |  |  |
| M±SD | 12.2±3.02 | 12.9±3.24 | 12.8±3.31 | 12.6±3.36 | 10.9±2.86 | 12.9±2.85 | 14.5±2.28 | 15.7±1.46 |

Education categories were 'none', 'other (non-university)', and 'university'. Townsend Deprivation Index scores capture areas with higher (positive score) or lower (negative score) material deprivation, relative to areas with average material deprivation (score of zero). Smoking categories were 'never', 'previous', or 'current'. Diet score categories were 'healthy' and 'unhealthy', defined according to dietary intake criteria for cardiometabolic health (see S3).

### Night and day light exposure predicts cardiovascular disease risk

Exposure to night light predicted higher risk of cardiovascular diseases, with clear dose-dependent relationships for each outcome (Figure 2; Table 2). Compared to people with dark nights (0-50^th^ light exposure percentiles), individuals with brighter nights had 1.11-1.12 (50-70^th^; adjusted-HR range for Models 1-3), 1.18-1.20 (70-90^th^), and 1.23-1.32 (90-100^th^) times higher risk of coronary artery disease. Similarly, those with brighter nights had 1.20-1.21 (50-70^th^), 1.27 (70-90^th^), and 1.42-1.47 (90-100^th^) times higher risk of myocardial infarction, and 1.15 (50-70^th^), 1.19-1.21 (70-90^th^), and 1.45-1.56 (90-100^th^ percentiles) times higher risk of heart failure. For those with the brightest nights (90-100^th^), we observed 1.28-1.32 times higher risk for atrial fibrillation and 1.28-1.30 times higher risk for stroke, compared to those with dark nights (0-50^th^ percentiles). Significant log-linear relationships of brighter night light with higher cardiovascular disease risks were observed for all outcomes (Table 3).

**Figure 2.**
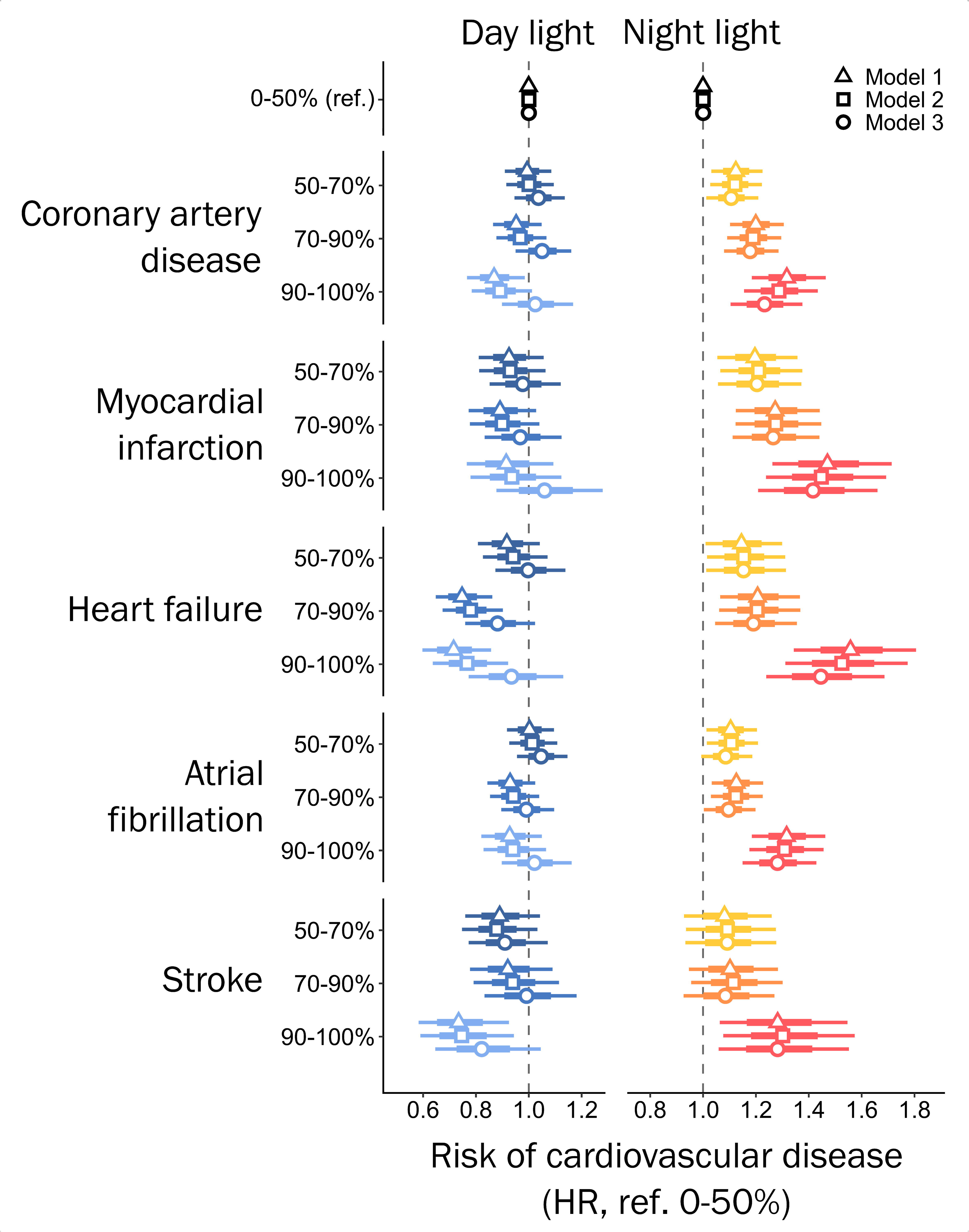
Hazard ratios for cardiovascular diseases within day and night light exposure percentile groups, adjusted for age, sex, and ethnicity (Model 1); additionally adjusted for education, employment, income, and deprivation (Model 2); and further adjusted for physical activity, smoking status, alcohol consumption, diet, and urbanicity (Model 3). Participants with the darkest environments (0-50^th^ percentiles) were the referent group for all models.

**Table 2.**
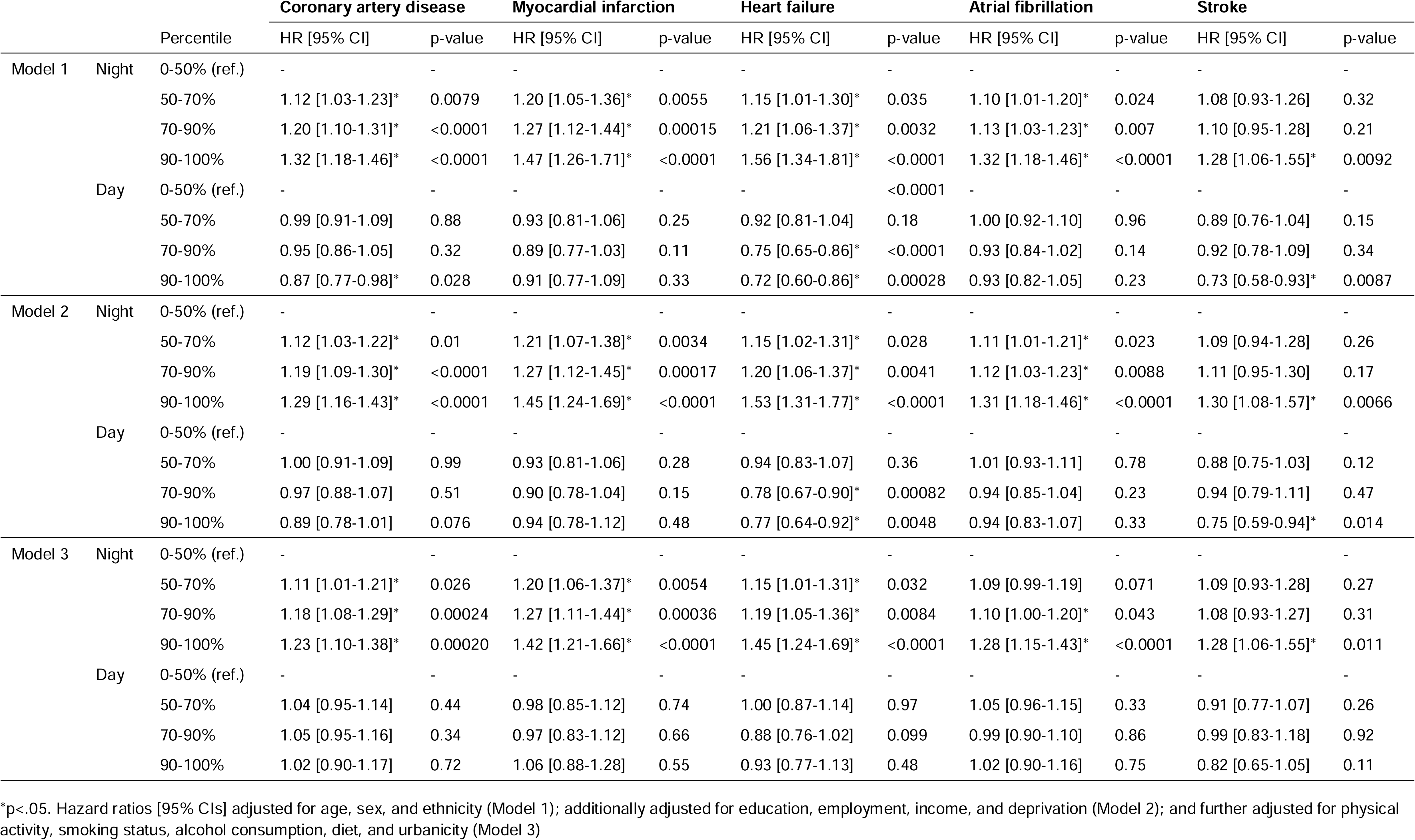
Risk of cardiovascular outcomes, according to light exposure percentile groups, across Models 1-3.

|  |  |  | Coronary artery disease |  | Myocardial infarction |  | Heart failure |  | Atrial fibrillation |  | Stroke |  |
| --- | --- | --- | --- | --- | --- | --- | --- | --- | --- | --- | --- | --- |
|  |  | Percentile | HR [95% CI] | p-value | HR [95% CI] | p-value | HR [95% CI] | p-value | HR [95% CI] | p-value | HR [95% CI] | p-value |
| Model 1 | Night | 0-50% (ref.) | - | - | - | - | - | - | - | - | - | - |
|  |  | 50-70% | 1.12 [1.03-1.23]* | 0.0079 | 1.20 [1.05-1.36]* | 0.0055 | 1.15 [1.01-1.30]* | 0.035 | 1.10 [1.01-1.20]* | 0.024 | 1.08 [0.93-1.26] | 0.32 |
|  |  | 70-90% | 1.20 [1.10-1.31]* | <0.0001 | 1.27 [1.12-1.44]* | 0.00015 | 1.21 [1.06-1.37]* | 0.0032 | 1.13 [1.03-1.23]* | 0.007 | 1.10 [0.95-1.28] | 0.21 |
|  |  | 90-100% | 1.32 [1.18-1.46]* | <0.0001 | 1.47 [1.26-1.71]* | <0.0001 | 1.56 [1.34-1.81]* | <0.0001 | 1.32 [1.18-1.46]* | <0.0001 | 1.28 [1.06-1.55]* | 0.0092 |
|  | Day | 0-50% (ref.) | - | - | - | - | - | <0.0001 | - | - | - | - |
|  |  | 50-70% | 0.99 [0.91-1.09] | 0.88 | 0.93 [0.81-1.06] | 0.25 | 0.92 [0.81-1.04] | 0.18 | 1.00 [0.92-1.10] | 0.96 | 0.89 [0.76-1.04] | 0.15 |
|  |  | 70-90% | 0.95 [0.86-1.05] | 0.32 | 0.89 [0.77-1.03] | 0.11 | 0.75 [0.65-0.86]* | <0.0001 | 0.93 [0.84-1.02] | 0.14 | 0.92 [0.78-1.09] | 0.34 |
|  |  | 90-100% | 0.87 [0.77-0.98]* | 0.028 | 0.91 [0.77-1.09] | 0.33 | 0.72 [0.60-0.86]* | 0.00028 | 0.93 [0.82-1.05] | 0.23 | 0.73 [0.58-0.93]* | 0.0087 |
| Model 2 | Night | 0-50% (ref.) | - | - | - | - | - | - | - | - | - | - |
|  |  | 50-70% | 1.12 [1.03-1.22]* | 0.01 | 1.21 [1.07-1.38]* | 0.0034 | 1.15 [1.02-1.31]* | 0.028 | 1.11 [1.01-1.21]* | 0.023 | 1.09 [0.94-1.28] | 0.26 |
|  |  | 70-90% | 1.19 [1.09-1.30]* | <0.0001 | 1.27 [1.12-1.45]* | 0.00017 | 1.20 [1.06-1.37]* | 0.0041 | 1.12 [1.03-1.23]* | 0.0088 | 1.11 [0.95-1.30] | 0.17 |
|  |  | 90-100% | 1.29 [1.16-1.43]* | <0.0001 | 1.45 [1.24-1.69]* | <0.0001 | 1.53 [1.31-1.77]* | <0.0001 | 1.31 [1.18-1.46]* | <0.0001 | 1.30 [1.08-1.57]* | 0.0066 |
|  | Day | 0-50% (ref.) | - | - | - | - | - | - | - | - | - | - |
|  |  | 50-70% | 1.00 [0.91-1.09] | 0.99 | 0.93 [0.81-1.06] | 0.28 | 0.94 [0.83-1.07] | 0.36 | 1.01 [0.93-1.11] | 0.78 | 0.88 [0.75-1.03] | 0.12 |
|  |  | 70-90% | 0.97 [0.88-1.07] | 0.51 | 0.90 [0.78-1.04] | 0.15 | 0.78 [0.67-0.90]* | 0.00082 | 0.94 [0.85-1.04] | 0.23 | 0.94 [0.79-1.11] | 0.47 |
|  |  | 90-100% | 0.89 [0.78-1.01] | 0.076 | 0.94 [0.78-1.12] | 0.48 | 0.77 [0.64-0.92]* | 0.0048 | 0.94 [0.83-1.07] | 0.33 | 0.75 [0.59-0.94]* | 0.014 |
| Model 3 | Night | 0-50% (ref.) | - | - | - | - | - | - | - | - | - | - |
|  |  | 50-70% | 1.11 [1.01-1.21]* | 0.026 | 1.20 [1.06-1.37]* | 0.0054 | 1.15 [1.01-1.31]* | 0.032 | 1.09 [0.99-1.19] | 0.071 | 1.09 [0.93-1.28] | 0.27 |
|  |  | 70-90% | 1.18 [1.08-1.29]* | 0.00024 | 1.27 [1.11-1.44]* | 0.00036 | 1.19 [1.05-1.36]* | 0.0084 | 1.10 [1.00-1.20]* | 0.043 | 1.08 [0.93-1.27] | 0.31 |
|  |  | 90-100% | 1.23 [1.10-1.38]* | 0.00020 | 1.42 [1.21-1.66]* | <0.0001 | 1.45 [1.24-1.69]* | <0.0001 | 1.28 [1.15-1.43]* | <0.0001 | 1.28 [1.06-1.55]* | 0.011 |
|  | Day | 0-50% (ref.) | - | - | - | - | - | - | - | - | - | - |
|  |  | 50-70% | 1.04 [0.95-1.14] | 0.44 | 0.98 [0.85-1.12] | 0.74 | 1.00 [0.87-1.14] | 0.97 | 1.05 [0.96-1.15] | 0.33 | 0.91 [0.77-1.07] | 0.26 |
|  |  | 70-90% | 1.05 [0.95-1.16] | 0.34 | 0.97 [0.83-1.12] | 0.66 | 0.88 [0.76-1.02] | 0.099 | 0.99 [0.90-1.10] | 0.86 | 0.99 [0.83-1.18] | 0.92 |
|  |  | 90-100% | 1.02 [0.90-1.17] | 0.72 | 1.06 [0.88-1.28] | 0.55 | 0.93 [0.77-1.13] | 0.48 | 1.02 [0.90-1.16] | 0.75 | 0.82 [0.65-1.05] | 0.11 |

\*p<.05. Hazard ratios [95% CIs] adjusted for age, sex, and ethnicity (Model 1); additionally adjusted for education, employment, income, and deprivation (Model 2); and further adjusted for physical activity, smoking status, alcohol consumption, diet, and urbanicity (Model 3)

**Table 3.**
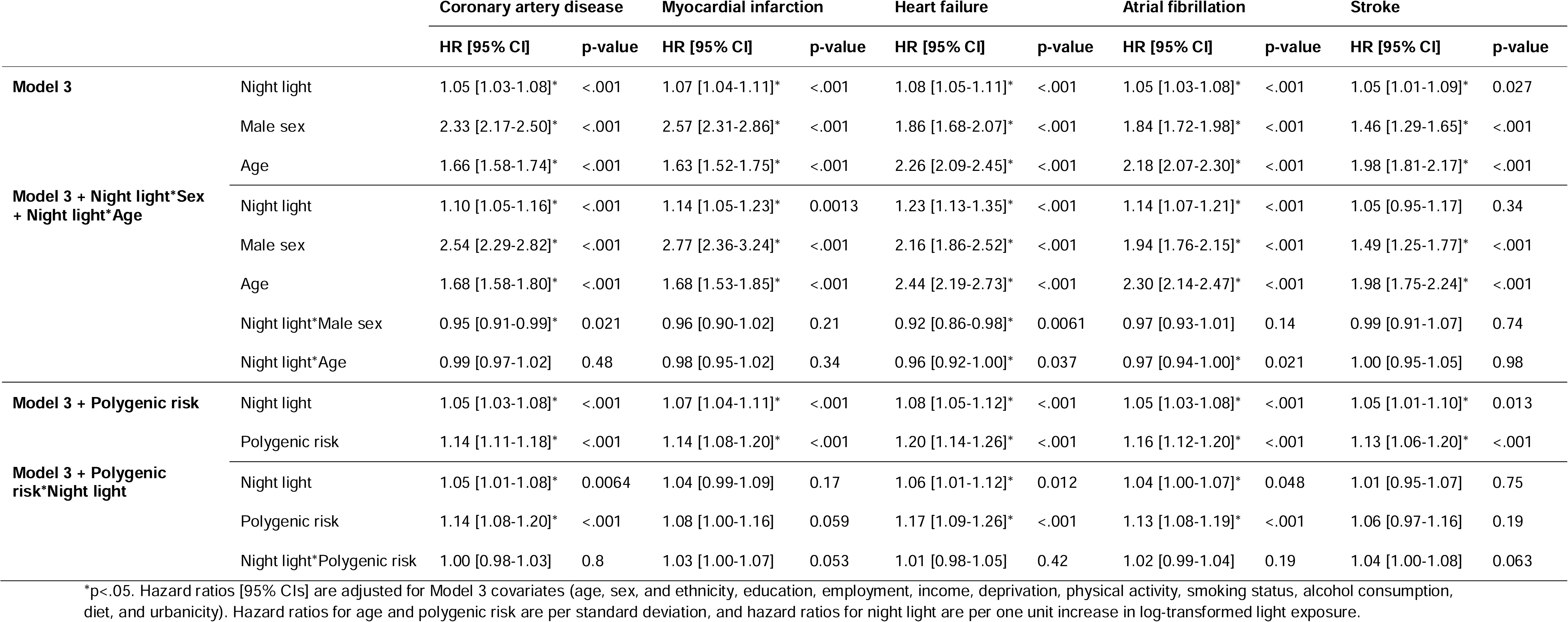
Cardiovascular risks related to standard deviation increases in night light exposure, including interactions with age, sex, and polygenic CVD risk.

Brighter light at night predicted higher cardiovascular disease risks following separate adjustments of Model 3 for pre-existing diabetes, hypertension, high BMI, high cholesterol ratio, short, long, or inefficient sleep, and exclusion of shift workers (Tables S5-10). The relationships of night light with cardiovascular risks were attenuated but remained statistically significant for all outcomes except stroke, which was attenuated below statistical significance after adjustment for short sleep and high cholesterol ratio.

Brighter day light exposure predicted lower risks of coronary artery disease, heart failure, and stroke in Models 1-2. Those with the brightest days (90-100^th^ percentiles) had 0.87-0.89, 0.72-0.77, and 0.73-0.75 times lower risk of coronary artery disease, heart failure, and stroke, respectively, compared to those with darker days (0-50^th^ percentiles), across Models 1-2. Trends towards brighter day light predicting lower risk of myocardial infarction and atrial fibrillation were observed, but these were not statistically significant. Brighter day light exposure did not predict risk of any cardiovascular disease in Model 3, which included additional adjustment for lifestyle factors (e.g., physical activity).

### Night light exposure and cardiovascular risk, according to age, sex, and genetic susceptibility

Higher risks of all cardiovascular outcomes were observed for males (aHR range: 1.46-2.57) compared to females, and with increasing age (aHR range: 1.63-2.26 per SD; Table 3). The strength of relationships of night light exposure with risks for heart failure, coronary artery disease, and atrial fibrillation varied according to participant age and/or sex (Table 3). Brighter night light was more strongly related to higher adjusted risk for heart failure in females (p-value for interaction=0.0061) and in younger individuals (p=0.037). Brighter night light diminished the protective effect of being female on risk of heart failure, such that females exposed to bright night light had similar heart failure risks to males exposed to bright night light (Figure S11). For risk of coronary artery disease, the relationship of brighter night light with higher adjusted risk was stronger in females than males (p=0.021; Figure S12). For risk of atrial fibrillation, the relationship of brighter night light with higher adjusted risk was stronger in younger individuals (p=0.021; Figure S13). The strength of relationships of brighter night light with risks for myocardial infarction did not vary according to age or sex.

Relationships of night light with higher cardiovascular risk were robust to including polygenic risk for CVD alongside model 3 covariates. No significant interactions of night light exposure with polygenic risk were observed for coronary artery disease, heart failure, atrial fibrillation, or stroke (Table 3).

## Discussion

Across ∼13 million hours of personal light exposure data, and ∼700,000 person-years of follow-up, individuals exposed to higher levels of personal night light had higher risks for incident coronary artery disease, myocardial infarction, heart failure, atrial fibrillation, and stroke. Importantly, the relationships of night light with cardiovascular disease risk were robust to adjustment for established cardiovascular risk factors including physical activity, diet, sleep, and genetic susceptibility. These findings support night light exposure as an important risk factor for adverse cardiovascular health.

The observed relationships of brighter personal night light with higher risk of cardiovascular diseases are consistent with previous studies of outdoor night light. We observed a 23-32% higher risk of coronary artery disease, and a 45-56% higher risk of myocardial infarction for people with the brightest nights (90-100th percentiles), compared to those with the darkest nights (0-50th percentiles). By contrast, a previous cohort study that used satellite data to define light exposure found that people with the brightest outdoor nights (top 20%) had a 7-23% greater risk of coronary heart disease, compared to those with the darkest outdoor nights (lowest 20%).^24^ We also observed a 28-30% higher risk of stroke for people with the brightest nights, compared to those with the darkest nights, whereas a previous satellite data study found a 26-43% higher risk of stroke for people with the brightest outdoor nights (top 25%), compared to those with darker outdoor nights (lowest 25%).^25^ Furthermore, we observed a 45-56% higher risk of heart failure, and a 28-32% higher risk of atrial fibrillation, for people with the brightest nights. To our knowledge, no previous large-scale studies have assessed relationships of light exposure with either heart failure or atrial fibrillation. Our study findings are also consistent with higher prevalence of cardiovascular disease risk factors in people with brighter nights, observed in smaller cohorts with objective light data.^26–28^ Finally, our findings are consistent with observed higher risks of adverse cardiovascular outcomes in rotating shift workers,^16–20^ a population that experiences frequent exposure to bright light during the biological night.

The observed higher risks of cardiovascular diseases in people with brighter nights may be explained by the disruptive effect of night light on circadian rhythms,^21–23^ as this can lead to dysregulation of various cardiovascular and metabolic mechanisms. First, circadian disruption is strongly implicated in impaired glucose tolerance^12,47^ and type 2 diabetes,^33,48,49^ which are significant risk factors for endothelial dysfunction and atherosclerosis. Second, circadian disruption may promote hypercoagulability,^9^ leading to higher risks of thromboembolic events and subsequent ischemia, particularly in people with atherosclerosis or atrial fibrillation. Third, circadian disruption can cause higher average 24-h blood pressure,^11,50,51^ potentially increasing the likelihood of vascular endothelial damage and myocardial hypertrophy. Finally, central circadian disruption may increase the likelihood of cardiac arrhythmia, due to conflicting inputs to the sinoatrial and atrioventricular nodes from the central circadian clock and cardiomyocyte clocks.^8^ Together, these mechanisms may explain the higher risks of coronary artery disease, myocardial infarction, heart failure, atrial fibrillation, and stroke with brighter night light exposure observed in this study.

Higher risks of cardiovascular diseases were observed for men, and older individuals, and the relationships of night light with heart failure, coronary artery disease, and atrial fibrillation differed according to age and/or sex. We observed stronger relationships of night light with risks of heart failure and coronary artery disease in women. These findings are consistent with previous research showing that exposure to shift work, which causes circadian disruption, predicts higher risk of heart failure in women compared to men.^18^ Greater sensitivity of the circadian system to bright light exposure has also been observed in women, compared to men.^52^ This may explain why women in this study had a greater increase in cardiovascular risk with bright nights as compared to men, such that the sex difference in these outcomes was less pronounced amongst those exposed to bright light. Stronger relationships of night light exposure with risk of heart failure and atrial fibrillation were observed for younger individuals. Brighter night light partially mitigated the protective effects of younger age for risk of these outcomes, such that young individuals with bright nights were observed to have risk profiles closer to those of older individuals. This finding may be partially attributable to attenuated circadian light sensitivity in older individuals.^53^

The dose-response relationships of brighter night light with higher risk of cardiovascular diseases were robust after accounting for individual genetic susceptibility for these diseases. This finding is important due to potential gene-environment confounding.^54,55^ For example, greater genetic susceptibility for cardiovascular diseases could influence both night light exposure behavior, and risk for developing a cardiovascular disease. We found that night light exposure is independent of polygenic risk for cardiovascular diseases, indicating that gene-environment correlations are unlikely to be driving the observed relationships.

Exposure to night light is a plausible proxy for sleep duration, which has been linked with risk of cardiovascular diseases.^56,57^ However, our findings indicate that relationships of brighter nights with higher risks of coronary artery disease, myocardial infarction, heart failure, and atrial fibrillation are independent of short, long, and inefficient sleep. The effects of the brightest night light on cardiovascular risks were attenuated but remained statistically significant following adjustment for short sleep duration, indicating that short sleep explained some, but not all, of the observed effects. These findings are consistent with experimental work demonstrating sleep-independent effects of light on circadian regulation of factors known to influence cardiovascular health, such as the secretion of glucagon-like peptide-1.^47^

### Strengths and Limitations

This study is the largest known examination of prospective links between personal light exposure and risks for incident cardiovascular diseases. Results were derived from ∼13 million hours of personal light exposure data from wrist-worn sensors, coupled with National Health Service records across a subsequent 9.5-year period, in ∼89,000 individuals. This cohort was well-characterized, including detailed sociodemographic and lifestyle information, objectively-measured sleep and physical activity, and genetic susceptibility for cardiovascular diseases. However, this study has several limitations. First, whether these findings generalize is not yet clear. The UKB cohort is predominantly white (97%), and over-represents individuals with higher education levels, higher income, women (57%), and those who are more likely to be healthy at the time of assessment.^58^ Second, longer-term light exposure measurements within each individual are needed to adequately capture the time-varying relationships of light exposure with cardiovascular disease risk, as opposed to the one week of light exposure collected in this study. Third, we note that some covariates included in analyses may be on causal pathways between night light exposure and cardiovascular risks (e.g., physical activity, diet), and therefore some models may be over-adjusted. Fourth, some covariates were collected prior to light tracking and may be subject to change over time. Finally, we note that these findings are observational, and do not capture the causal relationship of night light with cardiovascular disease risk. Long-term circadian-informed lighting interventions for individuals with high risk of adverse cardiometabolic outcomes are needed.

## Conclusions

Cardiovascular diseases are the leading cause of global morbidity and mortality.^59^ Current behavioral recommendations for preventing cardiovascular diseases include maintaining a healthy diet, attaining adequate physical activity, and avoiding alcohol and tobacco.^60^ This is the first study of personal light exposure patterns and incident cardiovascular diseases, establishing night light as an important new risk factor. Our findings demonstrate that, in addition to current recommendations, avoiding night light may be a promising approach for preventing cardiovascular diseases.

## Data Availability

All data used in the present work are available upon application to the UK Biobank team.

https://biobank.ndph.ox.ac.uk/showcase/

## Acknowledgements

This research was conducted using UK Biobank data (Project ID: 6818). We thank the UK Biobank team for developing and maintaining this resource, and the UK Biobank participants.

## Author Contributions

D.P.W. and A.C.B. had full access to all the data in the study and takes responsibility for the integrity of the data and the accuracy of the data analysis. D.P.W., A.C.B., A.J.K.P., and S.W.C. designed the research; D.P.W. and A.C.B. performed the research; D.P.W., A.C.B., A.J.K.P., and S.W.C. analysed the data; D.P.W. drafted the manuscript; D.P.W., A.C.B., J.M.L., M.K.R., R.S., F.A.J.L.S., A.J.K.P., and S.W.C. critically reviewed and contributed to the manuscript.

