## Supplementary Information for "Personal night light exposure predicts incidence of cardiovascular diseases in >88,000 individuals"

### Table S1. UK Biobank protocol documentation

| **Documentation** | **UK Biobank Website Link** |
| --- | --- |
| Invite to participate | https://biobank.ctsu.ox.ac.uk/crystal/refer.cgi?id=100253 |
| Participant instructions (Axivity AX3 device) | https://biobank.ndph.ox.ac.uk/showcase/refer.cgi?id=141141 |
| Collection and processing (Axivity AX3 device) | https://biobank.ndph.ox.ac.uk/showcase/refer.cgi?id=131600 |
| ‘First occurrence’ cardiovascular outcomes | https://biobank.ndph.ox.ac.uk/showcase/refer.cgi?id=593 |
| ‘Algorithmically defined’ cardiovascular outcomes | https://biobank.ndph.ox.ac.uk/showcase/refer.cgi?id=460 |
| Death register | https://biobank.ctsu.ox.ac.uk/crystal/refer.cgi?id=115559 |
| Ethics | https://www.ukbiobank.ac.uk/learn-more-about-uk-biobank/about-us/ethics |
| Reception and consent (assessment centre visit) | https://biobank.ndph.ox.ac.uk/showcase/refer.cgi?id=100230 |
|  | https://biobank.ndph.ox.ac.uk/showcase/ukb/docs/Reception.pdf |
| Physical measurements (assessment centre visit) | https://biobank.ndph.ox.ac.uk/showcase/refer.cgi?id=100225 |
|  | https://biobank.ndph.ox.ac.uk/showcase/refer.cgi?id=5636 |
|  | https://biobank.ndph.ox.ac.uk/showcase/refer.cgi?id=1227 |

### Section S2. Supplementary methods

**Cardiovascular outcomes**

Coronary artery disease was defined according to first occurrence outcomes and operations. First occurrences were defined according to ICD-10 codes for acute myocardial infarction (I21), subsequent myocardial infarction (I22), complications following myocardial infarction (I23), acute ischaemic heart disease (I24), and chronic ischaemic heart disease (I25). Operations were defined according to OPCS4 codes K40-46, K49, K50, and K75, or self-reported coronary angioplasty, coronary artery bypass, or triple heart bypass. Atrial fibrillation was defined according to first occurrence of ICD-10 code I48, or according to OPCS codes K62.1, K62.2, or K62.3.

**Day and night light exposure**

Factor analysis was applied to extract time windows where light exposure patterns exhibited clustered variance, according to the methodology reported in our previous work.^1^ In short, 48 half-hour light exposure bins, representing all clock times (e.g., 00:30 to 01:00), were used to extract night and day light factors. Varimax rotation with factor loading ≥0.5 was applied, and cumulative proportion of variance explained was 0.56. Factors were internally consistent (Cronbach’s α=.98 and α=.93, respectively), and weakly positively correlated (rs=0.10, p<.0001).

Night light exposure (00:30-06:00) was split into four percentile groups, with category boundaries as follows: <1.2 lx (0-50%); ≥1.2 & <6.2 lx (50-70%); ≥6.2 & < 48.3 lx (70-90%); and ≥48.3 lx (90-100%). Similarly, day light exposure (07:30-20:30) was split into four percentile groups, with category boundaries as follows: <991 lx (0-50%); ≥991 & <1750 lx (50-70%); ≥1750 & <3140 lx (70-90%); and ≥3140 lx (90-100%).

**Polygenic risk scores**

Polygenic risk scores (PRS) for CAD^2^, myocardial infarction^2^, heart failure^3^, atrial fibrillation^2^, and stroke^4^ was constructed using PRS-continuous shrinkage (PRS-CS).^5^ PRS–CS uses a Bayesian framework and a continuous shrinkage prior on SNP effect sizes while modelling linkage disequilibrium to improve polygenic prediction. These PRS were then scored in UK Biobank participants using PLINK 2^6^ as the weighted sum of the effect alleles, using the following formula:

$$S_{i}= \sum_{j=1}^{M} \hat{\beta}_{j}g_{ij}$$

where $S_{i}$is the polygenic score for individual *i*, $\hat{\beta}_{j}$ is the weighted additive effect of the effect allele at SNP *j*, and $g_{ij}$ is the genotype for individual *i* at SNP *j*.

**Genetic ancestry definition and principal components of ancestry**

European ancestry classification was completed using the Human Genome Diversity Project-1000 Genomes (HGDP-1KG) harmonized reference dataset.^7^ The HGDP-1KG is a high quality dataset of 4,094 whole genomes from labelled diverse continental populations. Principal components analysis (PCA) was performed on unrelated (KING kinship coefficient < 0∙125) individuals after pruning variants (500kb window, r2 = 0∙01) to extract the top 10 PCs of ancestry.^8-10^ We then projected individuals from the UK Biobank onto the HGDP-1KG PC space and trained a random forest classifier given continental ancestry labels from the HGDP-1KG cohort to assign ancestry to UK Biobank individuals based on their top 10 PC scores. The minimum random forest probability for assignment to a particular ancestry group was 0∙5 and we completed 20 iterations of this model. An individual was assigned to the European ancestry group and included in genetic cox models if 20/20 iterations assigned them to the European ancestry, otherwise individuals were excluded as non-European or admixed. Finally, PCA was completed within European UKB individuals to extract the top 5 PCs of ancestry for inclusion as population stratification covariates in genetic cox models.

**Model implementation**

Cox proportional hazards models were executed in R (version 4.4.1.), using the ‘survival’ package. The following is an example of model syntax:

*coxph(Surv(years, myocardial_infarction_diagnosis) ~ light_night + light_day + age + sex + ethnicity, data=data)*

Contrasts were set using the default ‘contr.treatment’ setting, for all categorical variables. The assumption of proportional hazards was assessed using the ‘cox.zph’ function.

Models assessing interactions of night light with age, sex, and genetic susceptibility as predictors of cardiovascular risks were implemented. In these models, night light was treated as a continuous variable, transformed according to the following equation:

*transformed_night_light = log(night_light + 1)*

Figures for each interaction model, generated figures represented marginal effects and were generated using the ‘sjPlot’ package (‘plot_model’ function, type = ‘pred’).

### Table S3. Covariates included in statistical analyses

| **Covariate** | **UKB ID(s)** | **Description** | **Model variable** |
| --- | --- | --- | --- |
| Age | 21003 | Obtained from NHS Primary Care Trust registries and confirmed with participants at assessment centre visit. | Continuous |
| Sex | 31 | Obtained from NHS Primary Care Trust registries and confirmed with participants at assessment centre visit. | Categorical: male, female |
| Ethnicity | 21000 | Ethnic group: white, mixed, Asian/Asian British, black/black British, Chinese, other, prefer not to say | Categorical: white, other |
| Education | 6138 | University, A Levels, O Levels, CSE, NVQ/HND/HNC, other, none, prefer not to say | Categorical: university (referent), other non-university (selection of any education category except university), none |
| Employment status | 6142 | Paid employment, unemployed, retired, home/family caretaker, unable to work, volunteer, student, other | Categorical: paid employment, other categories |
| Income | 738 | Yearly household income: <£18,000, £18,000-£29,900, £30,000-£51,900, £52,000-£100,000, >£100,000, do not know, prefer not to say | Categorical: <£18k, £18k-£29.9k, £30k-£51.9k, £52k-£100k, >£100k (referent), and unknown |
| Deprivation | 189 | Average home ownership, car ownership, household overcrowding, and employment rate of a participant’s local area. Derived using national census data at time of recruitment. | Continuous, included as recorded |
| Physical activity | 90012 | Accelerometer device average acceleration across one week of data collection | Continuous, included as recorded |
| Smoking status | 20116 | Smoking status: never, previous, current, prefer not to say | Categorical: current, previous, never (referent) |
| Alcohol consumption | 1558 | Alcohol intake frequency: daily, 3-4 times per week, 1-2 times per week, 1-3 times per month, special occasions only, never, prefer not to say | Continuous, days per week consuming alcohol: daily = 7, ‘3-4 times per week’ = 3.5, 1-2 times per week = 1.5, 1-3 times per month = 2*12/365.25*7, special occasions only = 1*12/365.25*7, never = 0 |
| Healthy diet | 1289, 1299, 1309, 1319, 1329, 1339, 1349, 1359, 1369, 1379, 1389, 1408, 1418, 1428, 1438, 1448, 1458, 1468, 2654, 3680, 6144 | Typical dietary intake. 10 nutritional intake criteria for cardiometabolic health were included, as reported previously^11^ | Categorical: healthy, unhealthy. Classified as healthy if ≥5 out of 10 dietary criteria were met. |
| Urbanicity | 20118 | Population density of participants’ local area, attained from the UK Office for National Statistics. | Categorical: urban (population ≥ 10,000), rural (population < 10,000) |
| Shift work | 826, 3426, 6142 | Participants’ work involves shift work or night shift work: never, sometimes, usually, always, do not know, prefer not to say | Categorical: shift-worker (‘sometimes’, ‘usually’ or 'always' for either ‘shift work’ or ‘night shift work’), non-shift worker (‘never/rarely’, or ‘unemployed’). |
| BMI | 21001 | Weight (kg) / height (m)^2^ | Categorial: high (BMI > 30), low (BMI ≤ 30) |
| Cholesterol ratio | 30760, 30780, 30870 | Blood biochemistry assays for high- and low-density lipoprotein, and triglycerides | Categorical: high (cholesterol ratio >3.75 for males or >3.00 for females), low (cholesterol ratio ≤3.75 for males or ≤3.00 for females). Calculated as cholesterol ratio = (HDL + LDL + 0.2*triglycerides)/HDL |
| Hypertension | 4080, 4079, 131286, 131288, 131290, 131292, 131294 | Physical measurement at assessment centre visit, two systolic and diastolic readings, averaged, or first occurrence of hypertension (ICD-10 codes I10-13, I15). | Categorical: high (systolic > 140, diastolic > 90, or diagnosed hypertension prior to light tracking), low (systolic ≤ 140, diastolic ≤ 90, or no diagnosed hypertension prior to light tracking) |
| Diabetes | 2443, 130706, 130708 | Diabetes diagnosed by a doctor: yes, no, do not know, prefer not to say, Diabetes diagnosed according to ICD-10 codes E10 or E11. | Categorical: diabetes, no diabetes |
| Photoperiod | N/A | Calculated from date of light tracking and coordinates of 53.4808° N, 2.2426° W (Manchester), using ‘getSunlightTimes()’ in the ‘suncalc’ package in R. | Continuous |

### Table S4. Participant characteristics for the total analysis sample, and for sub-groups without each cardiovascular outcome prior to light tracking

|  | **No exclusion** | **Coronary artery disease** | **Myocardial infarction** | **Heart failure** | **Atrial fibrillation** | **Stroke** |
| --- | --- | --- | --- | --- | --- | --- |
| Age |  |  |  |  |  |  |
| M±SD | 62.4±7.8 | 62.1±7.8 | 62.3±7.8 | 62.4±7.8 | 62.2±7.8 | 62.3±7.8 |
| Range | 43.5 to 79.0 | 43.5 to 79.0 | 43.5 to 79.0 | 43.5 to 79.0 | 43.5 to 79.0 | 43.5 to 79.0 |
| Sex (% male, N) | 43.1 (38321) | 41.6 (35205) | 42.3 (36748) | 42.9 (37836) | 42.4 (36558) | 42.9 (37661) |
| Ethnicity (% white, N) | 97.0 (85923) | 97.0 (81838) | 97.0 (83978) | 97.0 (85275) | 96.9 (83332) | 97.0 (84779) |
| Employment status (% employed, N) | 62.1 (54843) | 63.1 (53102) | 62.6 (53994) | 62.3 (54573) | 62.7 (53715) | 62.4 (54374) |
| Income |  |  |  |  |  |  |
| % <£18k, N | 13.0 (11502) | 12.6 (10615) | 12.8 (11057) | 13.0 (11350) | 12.9 (11059) | 12.9 (11240) |
| % £18-29.9k, N | 21.7 (19203) | 21.5 (18058) | 21.6 (18647) | 21.7 (19009) | 21.6 (18506) | 21.7 (18899) |
| % £30-51.9k, N | 26.0 (22912) | 26.1 (21946) | 26.0 (22461) | 26.0 (22782) | 26.0 (22256) | 26.0 (22663) |
| % £52-100k, N | 22.8 (20174) | 23.3 (19561) | 23.0 (19882) | 22.9 (20081) | 23.0 (19735) | 23.0 (19997) |
| % >£100k, N | 6.7 (5891) | 6.8 (5723) | 6.7 (5814) | 6.7 (5872) | 6.7 (5750) | 6.7 (5841) |
| Education |  |  |  |  |  |  |
| % other, N | 48.3 (42519) | 48.2 (40388) | 48.3 (41519) | 48.3 (42204) | 48.4 (41293) | 48.3 (41915) |
| % university/college, N | 43.4 (38176) | 43.9 (36843) | 43.7 (37556) | 43.4 (37952) | 43.5 (37123) | 43.5 (37775) |
| Townsend Deprivation Index |  |  |  |  |  |  |
| M±SD | -1.76±2.80 | -1.76±2.80 | -1.76±2.80 | -1.76±2.80 | -1.75±2.80 | -1.76±2.79 |
| Range | -6.26 to 10.5 | -6.26 to 10.5 | -6.26 to 10.5 | -6.26 to 10.5 | -6.26 to 10.5 | -6.26 to 10.5 |
| Smoking |  |  |  |  |  |  |
| % previous, N | 36.1 (31964) | 35.4 (29881) | 35.7 (30924) | 35.9 (31629) | 35.7 (30748) | 35.9 (31447) |
| % current, N | 6.8 (6057) | 6.8 (5713) | 6.8 (5845) | 6.8 (6005) | 6.9 (5911) | 6.8 (5956) |
| Alcohol (M±SD, days per week) | 2.98±2.50 | 2.97±2.50 | 2.98±2.50 | 2.98±2.50 | 2.97±2.49 | 2.98±2.50 |
| Urbanicity (% >10,000 population, N) | 84.1 (74033) | 84.1 (70530) | 84.1 (72377) | 84.1 (73486) | 84.2 (71899) | 84.1 (73089) |
| Physical activity |  |  |  |  |  |  |
| M±SD | 28.1±8.1 | 28.3±8.1 | 28.2±8.1 | 28.2±8.1 | 28.2±8.1 | 28.2±8.1 |
| Range | 4.8 to 69.4 | 4.8 to 69.4 | 4.8 to 69.4 | 4.83to 69.4 | 4.8 to 69.4 | 4.8 to 69.3 |
| Diet score (% healthy, N) | 25.8 (22258) | 25.7 (21194) | 25.8 (21779) | 25.8 (22096) | 25.7 (21544) | 25.8 (21977) |

### Table S5. Participant characteristics by light exposure percentiles, by day and night

|  | **Night light exposure percentile** | | | | **Day light exposure percentile** | | | |
| --- | --- | --- | --- | --- | --- | --- | --- | --- |
|  | **0-50%** | **50-70%** | **70-90%** | **90-100%** | **0-50%** | **50-70%** | **70-90%** | **90-100%** |
| BMI (% ≥30, N) | 16.9 (7481) | 19.8 (3519) | 22.2 (3934) | 26 (2305) | 19.7 (8740) | 19.9 (3523) | 19.2 (3404) | 17.7 (1572) |
| Diabetes (%, N) | 3.91 (1739) | 4.51 (802) | 5.36 (953) | 6.03 (536) | 4.7 (2089) | 4.64 (825) | 4.37 (777) | 3.81 (339) |
| Hypertension (%, N) | 26.3 (11707) | 26.7 (4739) | 27.6 (4910) | 30.8 (2736) | 27.3 (12141) | 26.7 (4741) | 26.8 (4774) | 27.4 (2436) |
| High cholesterol ratio* (%, N) | 65.8 (25125) | 66.8 (10233) | 66.3 (10127) | 68.1 (5253) | 66.5 (25415) | 66.9 (10193) | 66 (10125) | 65.1 (5005) |
| Sleep duration (% <6 h, N) | 12.1 (5189) | 20.6 (3536) | 27.2 (4665) | 40 (3418) | 20.1 (8553) | 19.4 (3335) | 19.2 (3295) | 18.9 (1625) |
| Sleep duration (% <9 h, N) | 1.56 (665) | 1.82 (312) | 0.671 (115) | 0.644 (55) | 1.48 (631) | 1.31 (225) | 1.11 (190) | 1.17 (101) |
| Sleep efficiency (range:0-100; M±SD) | 89.4±4.90 | 89.2±5.11 | 89.0±5.22 | 88.6±5.83 | 89.2±5.15 | 89.2±5.13 | 89.2±5.07 | 89.1±5.01 |
| Shift worker (%, N) | 6.61 (2918) | 8.84 (1560) | 9.81 (1730) | 10.1 (894) | 8.54 (3767) | 8.35 (1473) | 7.32 (1293) | 6.45 (569) |

*****High cholesterol ratio was defined as cholesterol ratio >3.75 for males or >3.00 for females

### Table S6. Relationships of day and night light with coronary artery disease, adjusted for pre-existing cardiometabolic health, sleep, and excluding shift workers

|  |  | **Percentile** | **Cases % (N)** | **HR [95% CI]** | **p-value** |
| --- | --- | --- | --- | --- | --- |
| Model 3 + BMI | Night | 0-50% (ref.) | 4.1 (1639) | - | - |
| N = 79908 |  | 50-70% | 4.39 (702) | 1.09 [1.00-1.19] | 0.053 |
|  |  | 70-90% | 4.73 (756) | 1.15 [1.06-1.26]* | 0.0013 |
|  |  | 90-100% | 5.16 (412) | 1.19 [1.06-1.33]* | 0.0021 |
|  | Day | 0-50% (ref.) | 4.21 (1684) | - | - |
|  |  | 50-70% | 4.44 (709) | 1.03 [0.94-1.13] | 0.47 |
|  |  | 70-90% | 4.56 (728) | 1.05 [0.95-1.16] | 0.38 |
|  |  | 90-100% | 4.86 (388) | 1.02 [0.90-1.17] | 0.72 |
| Model 3 + diabetes | Night | 0-50% (ref.) | 4.11 (1644) | - | - |
| N = 80058 |  | 50-70% | 4.4 (705) | 1.10 [1.00-1.20]* | 0.044 |
|  |  | 70-90% | 4.73 (758) | 1.16 [1.06-1.26]* | 0.001 |
|  |  | 90-100% | 5.17 (414) | 1.21 [1.08-1.35]* | 0.00084 |
|  | Day | 0-50% (ref.) | 4.23 (1692) | - | - |
|  |  | 50-70% | 4.43 (710) | 1.03 [0.94-1.13] | 0.48 |
|  |  | 70-90% | 4.57 (731) | 1.05 [0.95-1.16] | 0.31 |
|  |  | 90-100% | 4.85 (388) | 1.04 [0.91-1.18] | 0.6 |
| Model 3 + hypertension | Night | 0-50% (ref.) | 4.11 (1644) | - | - |
| N = 80058 |  | 50-70% | 4.4 (705) | 1.10 [1.00-1.20]* | 0.039 |
|  |  | 70-90% | 4.73 (758) | 1.17 [1.07-1.27]* | 0.00054 |
|  |  | 90-100% | 5.17 (414) | 1.20 [1.08-1.34]* | 0.001 |
|  | Day | 0-50% (ref.) | 4.23 (1692) | - | - |
|  |  | 50-70% | 4.43 (710) | 1.04 [0.95-1.14] | 0.4 |
|  |  | 70-90% | 4.57 (731) | 1.05 [0.95-1.16] | 0.34 |
|  |  | 90-100% | 4.85 (388) | 1.03 [0.91-1.18] | 0.64 |
| Model 3 + cholesterol ratio | Night | 0-50% (ref.) | 4.1 (1422) | - | - |
| N = 69288 |  | 50-70% | 4.5 (624) | 1.12 [1.02-1.23]* | 0.019 |
|  |  | 70-90% | 4.68 (648) | 1.16 [1.05-1.27]* | 0.0023 |
|  |  | 90-100% | 5.15 (357) | 1.20 [1.07-1.36]* | 0.0021 |
|  | Day | 0-50% (ref.) | 4.16 (1442) | - | - |
|  |  | 50-70% | 4.52 (627) | 1.08 [0.98-1.19] | 0.12 |
|  |  | 70-90% | 4.65 (644) | 1.09 [0.98-1.21] | 0.11 |
|  |  | 90-100% | 4.88 (338) | 1.05 [0.91-1.21] | 0.5 |
| Model 3 + short sleep duration | Night | 0-50% (ref.) | 4.1 (1582) | - | - |
| N = 77077 |  | 50-70% | 4.4 (679) | 1.08 [0.99-1.18] | 0.095 |
|  |  | 70-90% | 4.75 (733) | 1.13 [1.04-1.24]* | 0.0059 |
|  |  | 90-100% | 5.23 (403) | 1.16 [1.04-1.30]* | 0.01 |
|  | Day | 0-50% (ref.) | 4.21 (1621) | - | - |
|  |  | 50-70% | 4.48 (690) | 1.06 [0.96-1.16] | 0.24 |
|  |  | 70-90% | 4.59 (707) | 1.08 [0.97-1.19] | 0.16 |
|  |  | 90-100% | 4.92 (379) | 1.07 [0.93-1.22] | 0.34 |
| Model 3 + long sleep duration | Night | 0-50% (ref.) | 4.1 (1582) | - | - |
| N = 77077 |  | 50-70% | 4.4 (679) | 1.10 [1.01-1.21]* | 0.036 |
|  |  | 70-90% | 4.75 (733) | 1.18 [1.08-1.29]* | 0.00028 |
|  |  | 90-100% | 5.23 (403) | 1.25 [1.12-1.40]* | 0.0001 |
|  | Day | 0-50% (ref.) | 4.21 (1621) | - | - |
|  |  | 50-70% | 4.48 (690) | 1.05 [0.95-1.15] | 0.33 |
|  |  | 70-90% | 4.59 (707) | 1.06 [0.96-1.17] | 0.28 |
|  |  | 90-100% | 4.92 (379) | 1.04 [0.91-1.19] | 0.57 |
| Model 3 + sleep efficiency | Night | 0-50% (ref.) | 4.1 (1582) | - | - |
| N = 77077 |  | 50-70% | 4.4 (679) | 1.10 [1.00-1.20]* | 0.04 |
|  |  | 70-90% | 4.75 (733) | 1.17 [1.07-1.28]* | 0.00049 |
|  |  | 90-100% | 5.23 (403) | 1.23 [1.10-1.38]* | 0.0003 |
|  | Day | 0-50% (ref.) | 4.21 (1621) | - | - |
|  |  | 50-70% | 4.48 (690) | 1.05 [0.95-1.15] | 0.32 |
|  |  | 70-90% | 4.59 (707) | 1.06 [0.96-1.17] | 0.26 |
|  |  | 90-100% | 4.92 (379) | 1.04 [0.91-1.19] | 0.55 |
| Model 3, excluding shift workers | Night | 0-50% (ref.) | 4.2 (1543) | - | - |
| N = 73559 |  | 50-70% | 4.46 (656) | 1.12 [1.02-1.23]* | 0.016 |
|  |  | 70-90% | 4.61 (678) | 1.16 [1.05-1.27]* | 0.0019 |
|  |  | 90-100% | 5.15 (379) | 1.25 [1.11-1.40]* | 0.00015 |
|  | Day | 0-50% (ref.) | 4.23 (1557) | - | - |
|  |  | 50-70% | 4.49 (660) | 1.05 [0.95-1.15] | 0.36 |
|  |  | 70-90% | 4.62 (680) | 1.05 [0.95-1.16] | 0.37 |
|  |  | 90-100% | 4.88 (359) | 1.01 [0.88-1.15] | 0.91 |

Data are proportional hazards (95% CI). Model 3 was adjusted for: age, sex, ethnicity, yearly household income, area-level material deprivation, employment status, education, smoking status, alcohol consumption, healthy diet, physical activity, and urbanicity. * p<.05.

### Table S7. Relationships of day and night light with myocardial infarction, adjusted for pre-existing cardiometabolic health, sleep, and excluding shift workers

|  |  | **Percentile** | **Cases % (N)** | **HR [95% CI]** | **p-value** |
| --- | --- | --- | --- | --- | --- |
| Model 3 + BMI | Night | 0-50% (ref.) | 1.73 (709) | - | - |
| N = 81982 |  | 50-70% | 2.04 (335) | 1.19 [1.04-1.36]* | 0.0093 |
|  |  | 70-90% | 2.15 (352) | 1.24 [1.09-1.41]* | 0.0013 |
|  |  | 90-100% | 2.51 (206) | 1.38 [1.18-1.62]* | <0.0001 |
|  | Day | 0-50% (ref.) | 1.89 (776) | - | - |
|  |  | 50-70% | 1.9 (312) | 0.98 [0.85-1.12] | 0.72 |
|  |  | 70-90% | 1.95 (320) | 0.97 [0.83-1.13] | 0.68 |
|  |  | 90-100% | 2.37 (194) | 1.06 [0.88-1.28] | 0.53 |
| Model 3 + diabetes | Night | 0-50% (ref.) | 1.73 (710) | - | - |
| N = 82139 |  | 50-70% | 2.05 (336) | 1.19 [1.05-1.36]* | 0.0079 |
|  |  | 70-90% | 2.15 (354) | 1.25 [1.09-1.42]* | 0.00087 |
|  |  | 90-100% | 2.51 (206) | 1.39 [1.18-1.63]* | <0.0001 |
|  | Day | 0-50% (ref.) | 1.9 (779) | - | - |
|  |  | 50-70% | 1.91 (313) | 0.97 [0.85-1.12] | 0.71 |
|  |  | 70-90% | 1.95 (320) | 0.97 [0.84-1.13] | 0.7 |
|  |  | 90-100% | 2.36 (194) | 1.07 [0.89-1.29] | 0.47 |
| Model 3 + hypertension | Night | 0-50% (ref.) | 1.73 (710) | - | - |
| N = 82139 |  | 50-70% | 2.05 (336) | 1.20 [1.05-1.36]* | 0.0074 |
|  |  | 70-90% | 2.15 (354) | 1.25 [1.10-1.43]* | 0.00063 |
|  |  | 90-100% | 2.51 (206) | 1.38 [1.18-1.62]* | <0.0001 |
|  | Day | 0-50% (ref.) | 1.9 (779) | - | - |
|  |  | 50-70% | 1.91 (313) | 0.98 [0.85-1.12] | 0.78 |
|  |  | 70-90% | 1.95 (320) | 0.97 [0.83-1.12] | 0.66 |
|  |  | 90-100% | 2.36 (194) | 1.07 [0.88-1.29] | 0.5 |
| Model 3 + cholesterol ratio | Night | 0-50% (ref.) | 1.7 (603) | - | - |
| N = 71078 |  | 50-70% | 2.05 (291) | 1.21 [1.05-1.39]* | 0.008 |
|  |  | 70-90% | 2.12 (301) | 1.26 [1.09-1.45]* | 0.0015 |
|  |  | 90-100% | 2.45 (174) | 1.37 [1.15-1.63]* | 0.00035 |
|  | Day | 0-50% (ref.) | 1.84 (655) | - | - |
|  |  | 50-70% | 1.9 (270) | 1.01 [0.87-1.17] | 0.93 |
|  |  | 70-90% | 1.93 (275) | 0.99 [0.84-1.16] | 0.91 |
|  |  | 90-100% | 2.38 (169) | 1.10 [0.90-1.34] | 0.37 |
| Model 3 + short sleep duration | Night | 0-50% (ref.) | 1.73 (683) | - | - |
| N = 79079 |  | 50-70% | 2.05 (325) | 1.19 [1.04-1.35]* | 0.012 |
|  |  | 70-90% | 2.18 (345) | 1.24 [1.09-1.42]* | 0.0014 |
|  |  | 90-100% | 2.52 (199) | 1.35 [1.15-1.60]* | 0.00036 |
|  | Day | 0-50% (ref.) | 1.91 (755) | - | - |
|  |  | 50-70% | 1.91 (302) | 0.97 [0.85-1.12] | 0.69 |
|  |  | 70-90% | 1.93 (305) | 0.96 [0.82-1.11] | 0.56 |
|  |  | 90-100% | 2.4 (190) | 1.08 [0.89-1.30] | 0.45 |
| Model 3 + long sleep duration | Night | 0-50% (ref.) | 1.73 (683) | - | - |
| N = 79079 |  | 50-70% | 2.05 (325) | 1.20 [1.05-1.37]* | 0.0066 |
|  |  | 70-90% | 2.18 (345) | 1.27 [1.12-1.45]* | 0.00029 |
|  |  | 90-100% | 2.52 (199) | 1.42 [1.21-1.67]* | <0.0001 |
|  | Day | 0-50% (ref.) | 1.91 (755) | - | - |
|  |  | 50-70% | 1.91 (302) | 0.97 [0.84-1.11] | 0.62 |
|  |  | 70-90% | 1.93 (305) | 0.94 [0.81-1.10] | 0.45 |
|  |  | 90-100% | 2.4 (190) | 1.06 [0.87-1.28] | 0.58 |
| Model 3 + sleep efficiency | Night | 0-50% (ref.) | 1.73 (683) | - | - |
| N = 79079 |  | 50-70% | 2.05 (325) | 1.20 [1.05-1.37]* | 0.007 |
|  |  | 70-90% | 2.18 (345) | 1.27 [1.11-1.45]* | 0.00039 |
|  |  | 90-100% | 2.52 (199) | 1.40 [1.19-1.65]* | <0.0001 |
|  | Day | 0-50% (ref.) | 1.91 (755) | - | - |
|  |  | 50-70% | 1.91 (302) | 0.97 [0.84-1.11] | 0.62 |
|  |  | 70-90% | 1.93 (305) | 0.95 [0.81-1.10] | 0.47 |
|  |  | 90-100% | 2.4 (190) | 1.06 [0.87-1.28] | 0.56 |
| Model 3, excluding shift workers | Night | 0-50% (ref.) | 1.75 (661) | - | - |
| N = 75525 |  | 50-70% | 2.07 (312) | 1.23 [1.07-1.40]* | 0.0032 |
|  |  | 70-90% | 2.06 (311) | 1.23 [1.07-1.41]* | 0.0029 |
|  |  | 90-100% | 2.52 (190) | 1.45 [1.23-1.72]* | <0.0001 |
|  | Day | 0-50% (ref.) | 1.89 (714) | - | - |
|  |  | 50-70% | 1.89 (286) | 0.98 [0.84-1.13] | 0.73 |
|  |  | 70-90% | 1.93 (292) | 0.96 [0.82-1.12] | 0.6 |
|  |  | 90-100% | 2.41 (182) | 1.07 [0.88-1.30] | 0.51 |

Data are proportional hazards (95% CI). Model 3 was adjusted for: age, sex, ethnicity, yearly household income, area-level material deprivation, employment status, education, smoking status, alcohol consumption, healthy diet, physical activity, and urbanicity. * p<.05.

### Table S8. Relationships of day and night light with heart failure, adjusted for pre-existing cardiometabolic health, sleep, and excluding shift workers

|  |  | **Percentile** | **Cases % (N)** | **HR [95% CI]** | **p-value** |
| --- | --- | --- | --- | --- | --- |
| Model 3 + BMI | Night | 0-50% (ref.) | 1.83 (760) | - | - |
| N = 83245 |  | 50-70% | 1.95 (325) | 1.13 [0.99-1.28] | 0.074 |
|  |  | 70-90% | 2.03 (338) | 1.15 [1.01-1.31]* | 0.036 |
|  |  | 90-100% | 2.59 (216) | 1.36 [1.16-1.58]* | 0.00012 |
|  | Day | 0-50% (ref.) | 1.96 (814) | - | - |
|  |  | 50-70% | 2.04 (339) | 1.00 [0.87-1.14] | 0.97 |
|  |  | 70-90% | 1.83 (305) | 0.87 [0.75-1.01] | 0.069 |
|  |  | 90-100% | 2.17 (181) | 0.93 [0.77-1.13] | 0.46 |
| Model 3 + diabetes | Night | 0-50% (ref.) | 1.83 (763) | - | - |
| N = 83402 |  | 50-70% | 1.96 (327) | 1.14 [1.00-1.30] | 0.053 |
|  |  | 70-90% | 2.03 (339) | 1.16 [1.02-1.33]* | 0.022 |
|  |  | 90-100% | 2.61 (218) | 1.41 [1.21-1.64]* | <0.0001 |
|  | Day | 0-50% (ref.) | 1.96 (819) | - | - |
|  |  | 50-70% | 2.03 (339) | 0.99 [0.87-1.13] | 0.9 |
|  |  | 70-90% | 1.85 (308) | 0.88 [0.76-1.03] | 0.11 |
|  |  | 90-100% | 2.17 (181) | 0.95 [0.78-1.14] | 0.56 |
| Model 3 + hypertension | Night | 0-50% (ref.) | 1.83 (763) | - | - |
| N = 83402 |  | 50-70% | 1.96 (327) | 1.14 [1.00-1.30]* | 0.043 |
|  |  | 70-90% | 2.03 (339) | 1.17 [1.03-1.33]* | 0.017 |
|  |  | 90-100% | 2.61 (218) | 1.40 [1.20-1.63]* | <0.0001 |
|  | Day | 0-50% (ref.) | 1.96 (819) | - | - |
|  |  | 50-70% | 2.03 (339) | 1.00 [0.88-1.14] | 1 |
|  |  | 70-90% | 1.85 (308) | 0.88 [0.76-1.02] | 0.099 |
|  |  | 90-100% | 2.17 (181) | 0.95 [0.78-1.14] | 0.56 |
| Model 3 + cholesterol ratio | Night | 0-50% (ref.) | 1.81 (653) | - | - |
| N = 72171 |  | 50-70% | 2.02 (292) | 1.19 [1.03-1.36]* | 0.016 |
|  |  | 70-90% | 1.97 (284) | 1.16 [1.01-1.34]* | 0.039 |
|  |  | 90-100% | 2.54 (183) | 1.41 [1.19-1.66]* | <0.0001 |
|  | Day | 0-50% (ref.) | 1.94 (699) | - | - |
|  |  | 50-70% | 1.97 (285) | 0.98 [0.85-1.14] | 0.83 |
|  |  | 70-90% | 1.88 (271) | 0.90 [0.77-1.06] | 0.21 |
|  |  | 90-100% | 2.18 (157) | 0.94 [0.76-1.15] | 0.52 |
| Model 3 + short sleep duration | Night | 0-50% (ref.) | 1.83 (735) | - | <0.0001 |
| N = 80280 |  | 50-70% | 1.95 (313) | 1.11 [0.97-1.27] | 0.13 |
|  |  | 70-90% | 2.03 (326) | 1.13 [0.98-1.29] | 0.083 |
|  |  | 90-100% | 2.65 (213) | 1.34 [1.14-1.57]* | 0.00036 |
|  | Day | 0-50% (ref.) | 1.95 (783) | - | - |
|  |  | 50-70% | 2.06 (330) | 1.03 [0.90-1.17] | 0.72 |
|  |  | 70-90% | 1.85 (297) | 0.91 [0.78-1.06] | 0.22 |
|  |  | 90-100% | 2.2 (177) | 0.99 [0.82-1.20] | 0.92 |
| Model 3 + long sleep duration | Night | 0-50% (ref.) | 1.83 (735) | - | - |
| N = 80280 |  | 50-70% | 1.95 (313) | 1.13 [0.99-1.30] | 0.063 |
|  |  | 70-90% | 2.03 (326) | 1.18 [1.03-1.34]* | 0.016 |
|  |  | 90-100% | 2.65 (213) | 1.46 [1.24-1.70]* | <0.0001 |
|  | Day | 0-50% (ref.) | 1.95 (783) | - | - |
|  |  | 50-70% | 2.06 (330) | 1.01 [0.89-1.16] | 0.84 |
|  |  | 70-90% | 1.85 (297) | 0.89 [0.77-1.04] | 0.14 |
|  |  | 90-100% | 2.2 (177) | 0.96 [0.79-1.16] | 0.68 |
| Model 3 + sleep efficiency | Night | 0-50% (ref.) | 1.83 (735) | - | - |
| N = 80280 |  | 50-70% | 1.95 (313) | 1.13 [0.99-1.29] | 0.068 |
|  |  | 70-90% | 2.03 (326) | 1.17 [1.02-1.33]* | 0.023 |
|  |  | 90-100% | 2.65 (213) | 1.43 [1.22-1.67]* | <0.0001 |
|  | Day | 0-50% (ref.) | 1.95 (783) | - | - |
|  |  | 50-70% | 2.06 (330) | 1.02 [0.89-1.16] | 0.82 |
|  |  | 70-90% | 1.85 (297) | 0.90 [0.77-1.04] | 0.16 |
|  |  | 90-100% | 2.2 (177) | 0.97 [0.80-1.17] | 0.73 |
| Model 3, excluding shift workers | Night | 0-50% (ref.) | 1.9 (728) | - | - |
| N = 76716 |  | 50-70% | 2.03 (311) | 1.17 [1.02-1.34]* | 0.022 |
|  |  | 70-90% | 2.03 (311) | 1.17 [1.02-1.34]* | 0.021 |
|  |  | 90-100% | 2.54 (195) | 1.40 [1.19-1.65]* | <0.0001 |
|  | Day | 0-50% (ref.) | 2.01 (770) | - | - |
|  |  | 50-70% | 2.07 (317) | 0.99 [0.86-1.14] | 0.9 |
|  |  | 70-90% | 1.87 (287) | 0.87 [0.74-1.01] | 0.07 |
|  |  | 90-100% | 2.23 (171) | 0.92 [0.76-1.12] | 0.4 |

Data are proportional hazards (95% CI). Model 3 was adjusted for: age, sex, ethnicity, yearly household income, area-level material deprivation, employment status, education, smoking status, alcohol consumption, healthy diet, physical activity, and urbanicity. * p<.05.

### Table S9. Relationships of day and night light with atrial fibrillation, adjusted for pre-existing cardiometabolic health, sleep, and excluding shift workers

|  |  | **Percentile** | **Cases % (N)** | **HR [95% CI]** | **p-value** |
| --- | --- | --- | --- | --- | --- |
| Model 3 + BMI | Night | 0-50% (ref.) | 4.17 (1696) | - | - |
| N = 81382 |  | 50-70% | 4.21 (686) | 1.06 [0.97-1.16] | 0.2 |
|  |  | 70-90% | 4.29 (699) | 1.06 [0.97-1.16] | 0.18 |
|  |  | 90-100% | 5.16 (420) | 1.22 [1.09-1.36]* | 0.00039 |
|  | Day | 0-50% (ref.) | 4.1 (1668) | - | - |
|  |  | 50-70% | 4.42 (720) | 1.04 [0.95-1.14] | 0.38 |
|  |  | 70-90% | 4.34 (707) | 0.99 [0.89-1.09] | 0.83 |
|  |  | 90-100% | 4.99 (406) | 1.01 [0.89-1.15] | 0.83 |
| Model 3 + diabetes | Night | 0-50% (ref.) | 4.17 (1700) | - | - |
| N = 81537 |  | 50-70% | 4.24 (692) | 1.08 [0.99-1.18] | 0.096 |
|  |  | 70-90% | 4.3 (701) | 1.08 [0.99-1.18] | 0.078 |
|  |  | 90-100% | 5.18 (422) | 1.27 [1.13-1.41]* | <0.0001 |
|  | Day | 0-50% (ref.) | 4.12 (1678) | - | - |
|  |  | 50-70% | 4.43 (722) | 1.04 [0.95-1.14] | 0.35 |
|  |  | 70-90% | 4.34 (707) | 0.99 [0.90-1.10] | 0.89 |
|  |  | 90-100% | 5 (408) | 1.03 [0.90-1.17] | 0.68 |
| Model 3 + hypertension | Night | 0-50% (ref.) | 4.17 (1700) | - | - |
| N = 81537 |  | 50-70% | 4.24 (692) | 1.08 [0.99-1.18] | 0.089 |
|  |  | 70-90% | 4.3 (701) | 1.09 [0.99-1.19] | 0.068 |
|  |  | 90-100% | 5.18 (422) | 1.26 [1.13-1.40]* | <0.0001 |
|  | Day | 0-50% (ref.) | 4.12 (1678) | - | - |
|  |  | 50-70% | 4.43 (722) | 1.05 [0.96-1.15] | 0.3 |
|  |  | 70-90% | 4.34 (707) | 0.99 [0.90-1.10] | 0.87 |
|  |  | 90-100% | 5 (408) | 1.03 [0.90-1.17] | 0.66 |
| Model 3 + cholesterol ratio | Night | 0-50% (ref.) | 4.16 (1466) | - | - |
| N = 70548 |  | 50-70% | 4.35 (614) | 1.11 [1.01-1.22]* | 0.035 |
|  |  | 70-90% | 4.17 (588) | 1.06 [0.96-1.17] | 0.23 |
|  |  | 90-100% | 5.12 (361) | 1.26 [1.12-1.42]* | <0.0001 |
|  | Day | 0-50% (ref.) | 4.08 (1440) | - | - |
|  |  | 50-70% | 4.42 (624) | 1.06 [0.97-1.17] | 0.21 |
|  |  | 70-90% | 4.32 (609) | 1.00 [0.90-1.12] | 0.94 |
|  |  | 90-100% | 5.05 (356) | 1.05 [0.91-1.20] | 0.51 |
| Model 3 + short sleep duration | Night | 0-50% (ref.) | 4.17 (1637) | - | - |
| N = 78488 |  | 50-70% | 4.27 (671) | 1.08 [0.98-1.18] | 0.12 |
|  |  | 70-90% | 4.33 (679) | 1.08 [0.98-1.18] | 0.11 |
|  |  | 90-100% | 5.22 (410) | 1.25 [1.11-1.40]* | 0.00013 |
|  | Day | 0-50% (ref.) | 4.12 (1616) | - | - |
|  |  | 50-70% | 4.46 (700) | 1.05 [0.96-1.16] | 0.27 |
|  |  | 70-90% | 4.34 (681) | 1.00 [0.90-1.10] | 0.95 |
|  |  | 90-100% | 5.1 (400) | 1.05 [0.92-1.19] | 0.5 |
| Model 3 + long sleep duration | Night | 0-50% (ref.) | 4.17 (1637) | - | - |
| N = 78488 |  | 50-70% | 4.27 (671) | 1.09 [0.99-1.19] | 0.07 |
|  |  | 70-90% | 4.33 (679) | 1.10 [1.00-1.20]* | 0.039 |
|  |  | 90-100% | 5.22 (410) | 1.30 [1.16-1.45]* | <0.0001 |
|  | Day | 0-50% (ref.) | 4.12 (1616) | - | - |
|  |  | 50-70% | 4.46 (700) | 1.05 [0.96-1.15] | 0.31 |
|  |  | 70-90% | 4.34 (681) | 0.99 [0.89-1.09] | 0.82 |
|  |  | 90-100% | 5.1 (400) | 1.03 [0.91-1.18] | 0.64 |
| Model 3 + sleep efficiency | Night | 0-50% (ref.) | 4.17 (1637) | - | - |
| N = 78488 |  | 50-70% | 4.27 (671) | 1.09 [0.99-1.19] | 0.075 |
|  |  | 70-90% | 4.33 (679) | 1.09 [1.00-1.20] | 0.055 |
|  |  | 90-100% | 5.22 (410) | 1.28 [1.15-1.43]* | <0.0001 |
|  | Day | 0-50% (ref.) | 4.12 (1616) | - | - |
|  |  | 50-70% | 4.46 (700) | 1.05 [0.96-1.15] | 0.31 |
|  |  | 70-90% | 4.34 (681) | 0.99 [0.89-1.10] | 0.84 |
|  |  | 90-100% | 5.1 (400) | 1.04 [0.91-1.18] | 0.61 |
| Model 3, excluding shift workers | Night | 0-50% (ref.) | 4.31 (1616) | - | - |
| N = 74943 |  | 50-70% | 4.39 (658) | 1.10 [1.01-1.21]* | 0.034 |
|  |  | 70-90% | 4.38 (656) | 1.10 [1.01-1.21]* | 0.036 |
|  |  | 90-100% | 5.11 (383) | 1.26 [1.13-1.41]* | <0.0001 |
|  | Day | 0-50% (ref.) | 4.24 (1587) | - | - |
|  |  | 50-70% | 4.47 (670) | 1.03 [0.94-1.13] | 0.57 |
|  |  | 70-90% | 4.48 (672) | 0.99 [0.89-1.10] | 0.85 |
|  |  | 90-100% | 5.12 (384) | 1.00 [0.88-1.15] | 0.97 |

Data are proportional hazards (95% CI). Model 3 was adjusted for: age, sex, ethnicity, yearly household income, area-level material deprivation, employment status, education, smoking status, alcohol consumption, healthy diet, physical activity, and urbanicity. * p<.05.

### Table S10. Relationships of day and night light with stroke, adjusted for pre-existing cardiometabolic health, sleep, and excluding shift workers

|  |  | **Percentile** | **Cases % (N)** | **HR [95% CI]** | **p-value** |
| --- | --- | --- | --- | --- | --- |
| Model 3 + BMI | Night | 0-50% (ref.) | 1.33 (549) | - | - |
| N = 82773 |  | 50-70% | 1.33 (220) | 1.08 [0.93-1.27] | 0.32 |
|  |  | 70-90% | 1.33 (221) | 1.08 [0.92-1.27] | 0.33 |
|  |  | 90-100% | 1.66 (137) | 1.28 [1.06-1.56]* | 0.01 |
|  | Day | 0-50% (ref.) | 1.39 (574) | - | - |
|  |  | 50-70% | 1.29 (213) | 0.90 [0.77-1.07] | 0.23 |
|  |  | 70-90% | 1.41 (234) | 0.98 [0.83-1.17] | 0.86 |
|  |  | 90-100% | 1.28 (106) | 0.81 [0.64-1.03] | 0.088 |
| Model 3 + diabetes | Night | 0-50% (ref.) | 1.33 (550) | - | - |
| N = 82931 |  | 50-70% | 1.34 (222) | 1.09 [0.93-1.27] | 0.3 |
|  |  | 70-90% | 1.34 (222) | 1.08 [0.92-1.26] | 0.37 |
|  |  | 90-100% | 1.65 (137) | 1.27 [1.05-1.54]* | 0.014 |
|  | Day | 0-50% (ref.) | 1.39 (575) | - | - |
|  |  | 50-70% | 1.29 (214) | 0.91 [0.77-1.07] | 0.25 |
|  |  | 70-90% | 1.42 (235) | 0.99 [0.83-1.18] | 0.94 |
|  |  | 90-100% | 1.29 (107) | 0.83 [0.65-1.05] | 0.12 |
| Model 3 + hypertension | Night | 0-50% (ref.) | 1.33 (550) | - | - |
| N = 82931 |  | 50-70% | 1.34 (222) | 1.09 [0.93-1.27] | 0.29 |
|  |  | 70-90% | 1.34 (222) | 1.08 [0.92-1.26] | 0.36 |
|  |  | 90-100% | 1.65 (137) | 1.26 [1.04-1.53]* | 0.016 |
|  | Day | 0-50% (ref.) | 1.39 (575) | - | - |
|  |  | 50-70% | 1.29 (214) | 0.91 [0.77-1.07] | 0.27 |
|  |  | 70-90% | 1.42 (235) | 0.99 [0.83-1.18] | 0.93 |
|  |  | 90-100% | 1.29 (107) | 0.83 [0.65-1.05] | 0.12 |
| Model 3 + cholesterol ratio | Night | 0-50% (ref.) | 1.38 (497) | - | - |
| N = 71769 |  | 50-70% | 1.38 (198) | 1.06 [0.9-1.25] | 0.47 |
|  |  | 70-90% | 1.36 (195) | 1.05 [0.89-1.24] | 0.55 |
|  |  | 90-100% | 1.62 (116) | 1.18 [0.96-1.46] | 0.11 |
|  | Day | 0-50% (ref.) | 1.43 (513) | - | - |
|  |  | 50-70% | 1.32 (189) | 0.90 [0.76-1.07] | 0.23 |
|  |  | 70-90% | 1.44 (206) | 0.97 [0.81-1.17] | 0.75 |
|  |  | 90-100% | 1.37 (98) | 0.84 [0.65-1.08] | 0.17 |
| Model 3 + short sleep duration | Night | 0-50% (ref.) | 1.32 (528) | - | - |
| N = 79829 |  | 50-70% | 1.33 (212) | 1.06 [0.9-1.24] | 0.48 |
|  |  | 70-90% | 1.37 (218) | 1.07 [0.91-1.26] | 0.42 |
|  |  | 90-100% | 1.64 (131) | 1.20 [0.98-1.47] | 0.074 |
|  | Day | 0-50% (ref.) | 1.38 (550) | - | - |
|  |  | 50-70% | 1.31 (209) | 0.93 [0.78-1.09] | 0.36 |
|  |  | 70-90% | 1.43 (228) | 1.01 [0.84-1.2] | 0.95 |
|  |  | 90-100% | 1.28 (102) | 0.82 [0.64-1.05] | 0.12 |
| Model 3 + long sleep duration | Night | 0-50% (ref.) | 1.32 (528) | - | - |
| N = 79829 |  | 50-70% | 1.33 (212) | 1.08 [0.92-1.27] | 0.34 |
|  |  | 70-90% | 1.37 (218) | 1.12 [0.96-1.32] | 0.16 |
|  |  | 90-100% | 1.64 (131) | 1.30 [1.07-1.58]* | 0.0089 |
|  | Day | 0-50% (ref.) | 1.38 (550) | - | - |
|  |  | 50-70% | 1.31 (209) | 0.92 [0.78-1.09] | 0.32 |
|  |  | 70-90% | 1.43 (228) | 0.99 [0.83-1.18] | 0.91 |
|  |  | 90-100% | 1.28 (102) | 0.80 [0.62-1.02] | 0.072 |
| Model 3 + sleep efficiency | Night | 0-50% (ref.) | 1.32 (528) | - | - |
| N = 79829 |  | 50-70% | 1.33 (212) | 1.08 [0.92-1.27] | 0.35 |
|  |  | 70-90% | 1.37 (218) | 1.11 [0.94-1.3] | 0.21 |
|  |  | 90-100% | 1.64 (131) | 1.28 [1.05-1.56]* | 0.013 |
|  | Day | 0-50% (ref.) | 1.38 (550) | - | - |
|  |  | 50-70% | 1.31 (209) | 0.92 [0.78-1.08] | 0.32 |
|  |  | 70-90% | 1.43 (228) | 0.99 [0.83-1.18] | 0.92 |
|  |  | 90-100% | 1.28 (102) | 0.80 [0.63-1.03] | 0.079 |
| Model 3, excluding shift workers | Night | 0-50% (ref.) | 1.38 (525) | - | - |
| N = 76261 |  | 50-70% | 1.37 (209) | 1.09 [0.93-1.29] | 0.27 |
|  |  | 70-90% | 1.39 (212) | 1.11 [0.95-1.31] | 0.19 |
|  |  | 90-100% | 1.6 (122) | 1.24 [1.01-1.52]* | 0.036 |
|  | Day | 0-50% (ref.) | 1.43 (545) | - | - |
|  |  | 50-70% | 1.32 (201) | 0.89 [0.76-1.06] | 0.2 |
|  |  | 70-90% | 1.46 (222) | 0.97 [0.81-1.16] | 0.72 |
|  |  | 90-100% | 1.31 (100) | 0.78 [0.61-1] | 0.051 |

Data are proportional hazards (95% CI). Model 3 was adjusted for: age, sex, ethnicity, yearly household income, area-level material deprivation, employment status, education, smoking status, alcohol consumption, healthy diet, physical activity, and urbanicity. * p<.05.

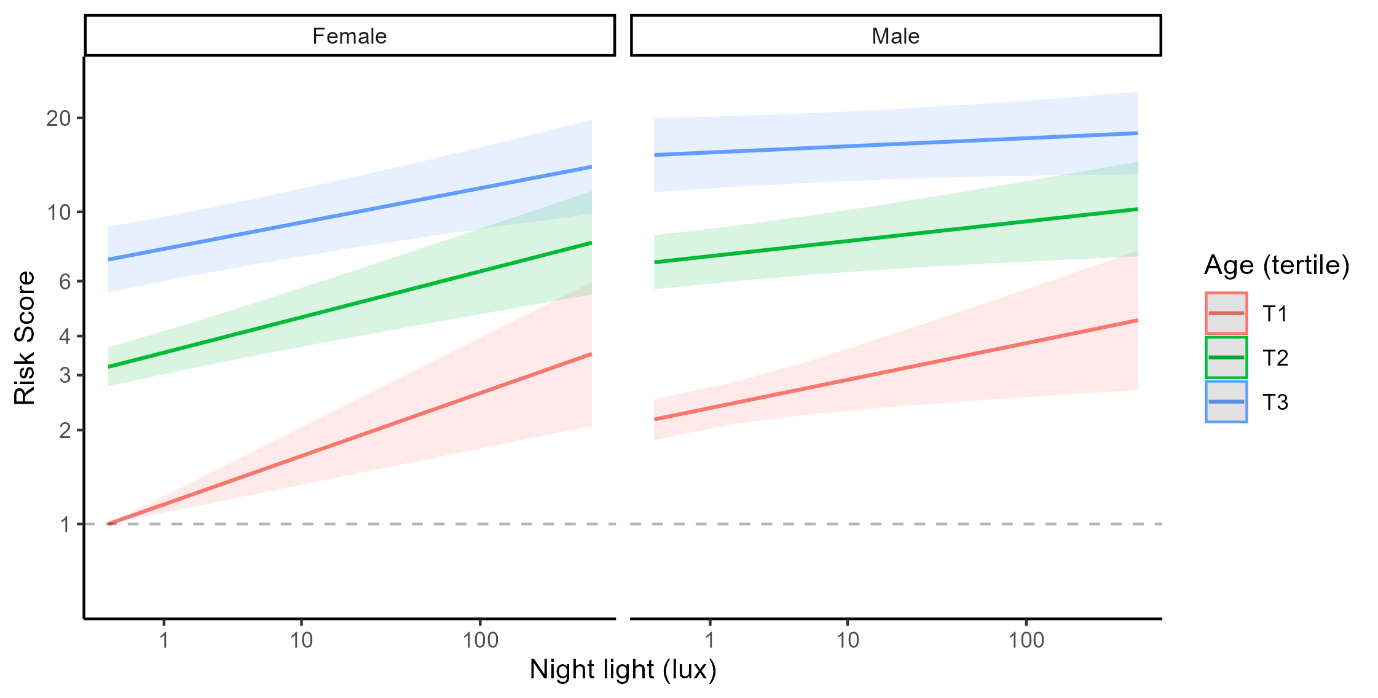

### Figure S11. Relationship of night light exposure with risk of heart failure, according to participant age and sex

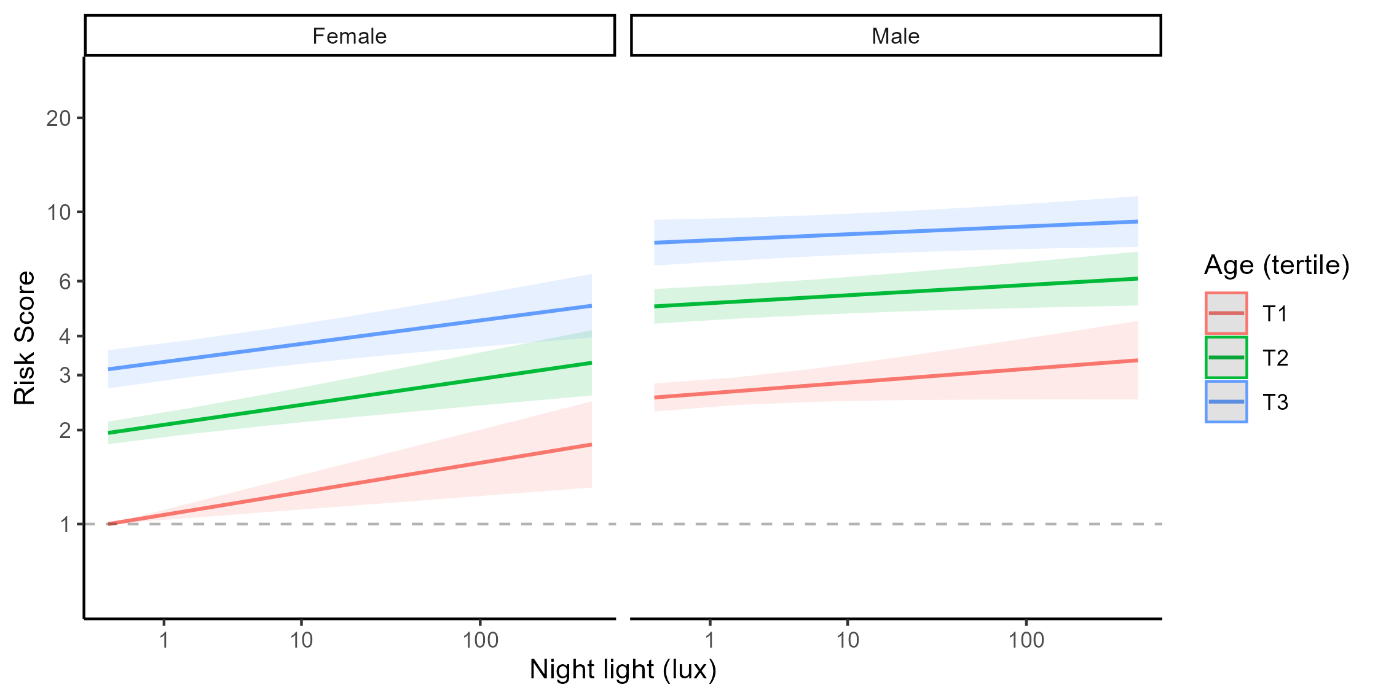

### Figure S12. Relationship of night light exposure with risk of coronary artery disease, according to participant age and sex

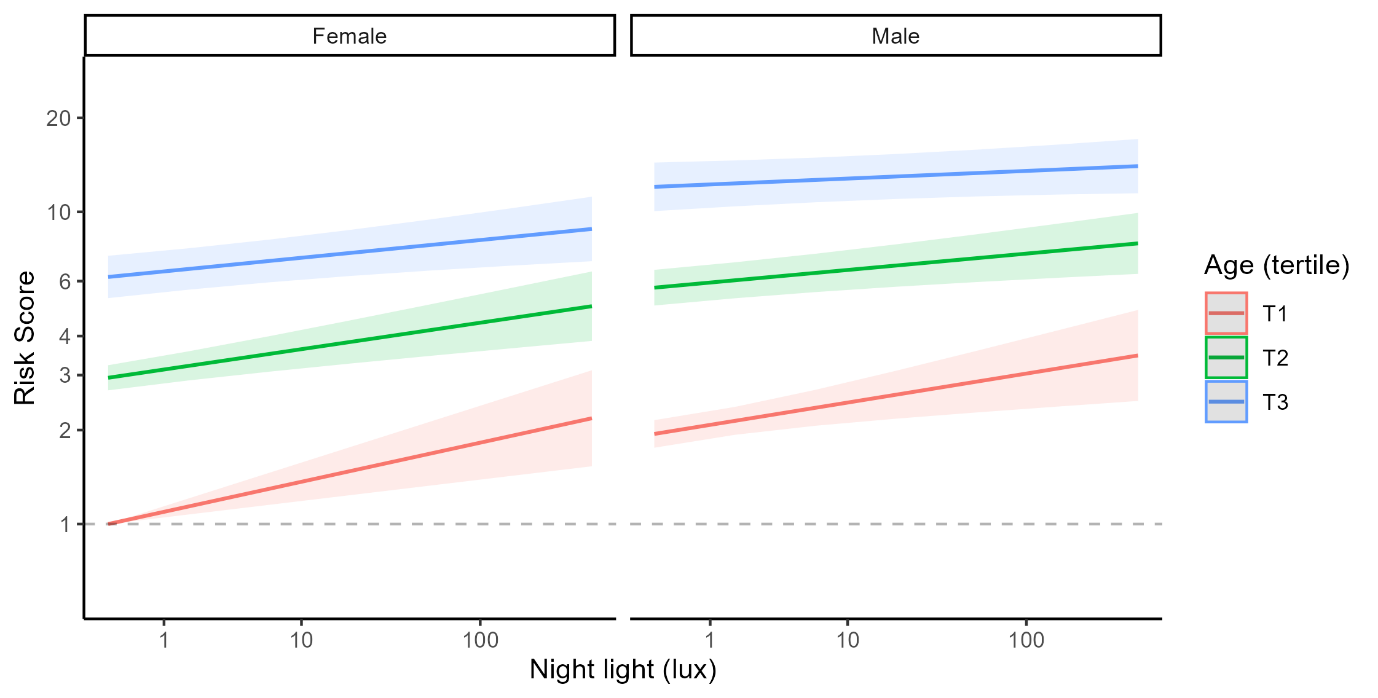

### Figure S13. Relationship of night light exposure with risk of atrial fibrillation, according to participant age and sex

### Section S14. STROBE Statement

STROBE Statement—Checklist of items that should be included in reports of *cohort studies*

|  | Item No | Recommendation | Manuscript Section |
| --- | --- | --- | --- |
| Title and abstract | 1 | (*a*) Indicate the study’s design with a commonly used term in the title or the abstract | - Abstract |
|  |  | (*b*) Provide in the abstract an informative and balanced summary of what was done and what was found | - Abstract |
| Introduction | | |  |
| Background/rationale | 2 | Explain the scientific background and rationale for the investigation being reported | - Introduction |
| Objectives | 3 | State specific objectives, including any prespecified hypotheses | - Introduction |
| Methods | | |  |
| Study design | 4 | Present key elements of study design early in the paper | - Abstract; Introduction; Methods |
| Setting | 5 | Describe the setting, locations, and relevant dates, including periods of recruitment, exposure, follow-up, and data collection | - Abstract; Introduction; Methods |
| Participants | 6 | (*a*) Give the eligibility criteria, and the sources and methods of selection of participants. Describe methods of follow-up | - Methods: Overview; Figure 1 |
|  |  | (*b*) For matched studies, give matching criteria and number of exposed and unexposed | - N/A |
| Variables | 7 | Clearly define all outcomes, exposures, predictors, potential confounders, and effect modifiers. Give diagnostic criteria, if applicable | - Methods; Supplementary S2 |
| Data sources/ measurement | 8* | For each variable of interest, give sources of data and details of methods of assessment (measurement). Describe comparability of assessment methods if there is more than one group | - Methods; Supplementary S3 |
| Bias | 9 | Describe any efforts to address potential sources of bias | - Methods |
| Study size | 10 | Explain how the study size was arrived at | - Methods; Figure 1 |
| Quantitative variables | 11 | Explain how quantitative variables were handled in the analyses. If applicable, describe which groupings were chosen and why | - Methods; Supplementary S3 |
| Statistical methods | 12 | (*a*) Describe all statistical methods, including those used to control for confounding | - Methods; Supplementary S2 |
|  |  | (*b*) Describe any methods used to examine subgroups and interactions | - Methods; Supplementary S2 |
|  |  | (*c*) Explain how missing data were addressed | - Methods; Supplementary S4 |
|  |  | (*d*) If applicable, explain how loss to follow-up was addressed | - N/A |
|  |  | (*e*) Describe any sensitivity analyses | - Methods; Supplementary S5-13 |
| Results | | |  |
| Participants | 13* | (a) Report numbers of individuals at each stage of study—e.g., numbers potentially eligible, examined for eligibility, confirmed eligible, included in the study, completing follow-up, and analysed | - Figure 1 |
|  |  | (b) Give reasons for non-participation at each stage | - Figure 1 |
|  |  | (c) Consider use of a flow diagram | - Figure 1 |
| Descriptive data | 14* | (a) Give characteristics of study participants (e.g., demographic, clinical, social) and information on exposures and potential confounders | - Table 1; Supplementary S4 |
|  |  | (b) Indicate number of participants with missing data for each variable of interest | - Figure 1 |
|  |  | (c) Summarise follow-up time (e.g., average and total amount) | - Results |
| Outcome data | 15* | Report numbers of outcome events or summary measures over time | - Table 1 |
| Main results | 16 | (*a*) Give unadjusted estimates and, if applicable, confounder-adjusted estimates and their precision (e.g., 95% confidence interval). Make clear which confounders were adjusted for and why they were included | - Results |
|  |  | (*b*) Report category boundaries when continuous variables were categorized | - Supplementary S2 |
|  |  | (*c*) If relevant, consider translating estimates of relative risk into absolute risk for a meaningful time period | - N/A |
| Other analyses | 17 | Report other analyses done—e.g., analyses of subgroups and interactions, and sensitivity analyses | - Results; Supplementary S11-S13 |
| Discussion | | |  |
| Key results | 18 | Summarise key results with reference to study objectives | - Discussion paragraphs 1,2 |
| Limitations | 19 | Discuss limitations of the study, taking into account sources of potential bias or imprecision. Discuss both direction and magnitude of any potential bias | - Discussion: Strengths and limitations |
| Interpretation | 20 | Give a cautious overall interpretation of results considering objectives, limitations, multiplicity of analyses, results from similar studies, and other relevant evidence | - Discussion: paragraphs 1, 3, Conclusions |
| Generalisability | 21 | Discuss the generalisability (external validity) of the study results | - Discussion: Strengths and limitations |
| Other information | | |  |
| Funding | 22 | Give the source of funding and the role of the funders for the present study and, if applicable, for the original study on which the present article is based | - Acknowledgements |
